# Application of deep learning and explainable AI-supported medical decision-making for facial phenotyping in genetic syndromes

**DOI:** 10.1101/2025.06.08.25328588

**Authors:** Ömer Sümer, Tobias Huber, Jiayi Cheng, Dat Duong, Suzanna E. Ledgister Hanchard, Cristina Conati, Elisabeth André, Benjamin D Solomon, Rebekah L. Waikel

## Abstract

**Objective:** To assess whether saliency-based explainable artificial intelligence (XAI) can improve diagnostic accuracy, confidence, and perceived usefulness for recognition of genetic conditions based on facial images.

**Materials and methods:** Forty-four medical geneticists, divided into AI-only (n=23) and XAI-supported (n=21) groups, assessed eighteen images of individuals with and without genetic conditions. Geneticists’ diagnostic accuracy and confidence were recorded before and after viewing an AI classifier’s prediction probability, with or without XAI explanations. Mediation analyses were conducted to interpret geneticist behavior.

**Results:** AI-only and XAI-support improved geneticist accuracy for correct AI classifications, with mean improvements of 0.20±0.13 (AI-only) and 0.19±0.13 (XAI). Incorrect AI classifications decreased accuracy in both groups: −0.20±0.22 (AI-only) and −0.21±0.23 (XAI). Average confidence increased with correct classification: 0.30±0.26 (AI-only) and 0.38±0.71 (XAI) and decreased with incorrect classification: −0.14±0.64 (AI-only) and - 0.14±0.76 (XAI). AI prediction probability were generally considered useful (0.58±1.1 (AI-only) and 0.72±1.3 (XAI)); XAI explanations were viewed less favorably: −0.14±1.3 (saliency maps); −0.19±1.3 (region relevance scores). For incorrect AI classifications, there was a negative correlation between accuracy improvement and perceived AI usefulness (Spearman’s Rho −0.31 (AI-only) and −0.40 (XAI)). Mediation analyses demonstrated that when AI is correct (without XAI), there was a significant mediated effect (0.583, 95% CI [0.044, 1.144]) by the model probability between user confidence and decision to follow AI.

**Conclusions:** The lack of accuracy or confidence improvements together with additional qualitative responses indicate that participants did not integrate saliency-based XAI into their decisions. AI prediction probability had a greater impact on participants’ decision making.

## INTRODUCTION

### Background and Significance

While rare diseases collectively affect nearly 1 in 10 individuals, individual rare diseases affect fewer than 1 in 2000 (per European definitions). Most rare diseases have genetic causes, and many are extremely uncommon, making diagnosis challenging. Artificial intelligence (AI) driven tools, especially Deep learning (DL), have advanced not only biomedical research in general but can also be used to analyze facial images and provide differential diagnoses by detecting distinctive features of these rare diseases. [1] [2]

Interestingly, previous studies have suggested that AI tools may not improve human diagnostic reasoning; for instance, although large language models (LLMs) outperform physicians, LLMs fail to enhance decision-making.[3] In another a systematic review, results of AI-based clinical decision support systems have been mixed; the authors found sparse evidence that the use of such systems is associated with improved clinician performance at diagnostic tasks.[4 5] Such examples underscore the complexity of human-AI collaboration, difficulties in assessing effects, and the need to investigate the interplay between AI performance, clinician trust, and diagnostic behavior in many instances,[6] including facial phenotyping in medical genetics.

Explainable AI (XAI) methods are emerging to validate that AI-generated insights are interpretable, trustworthy, and verifiable by medical professionals. XAI is comprised of many different approaches; for example, GAN counterfactual,[7] self-reflection in LLM,[8] Shapely values in decision tree,[9] and saliency maps in image classification.[10]

Given that our objectives involve image classification, we focused on saliency methods. Specifically, we evaluated the usefulness of saliency maps produced by a DL classifier to predict genetic conditions based on human faces. We used these saliency maps along with a novel approach, region relevance scores.[11] In this study, we use the term XAI to encompass the saliency maps and the region relevance scores.

In our previous work, we found that when human experts assess facial images in this way, they primarily pay attention to three main areas of interest (AOI): nose, eyes, and mouth.[12] Using these three main AOIs, our region relevance scores quantitatively summarize the output of a saliency map, indicating which of the 3 AOIs are contributing the most to the model predication probabilities/syndrome classification. Understanding how medical geneticists assess facial dysmorphology and perceive AI-driven outputs is critical for improving human-AI collaboration. These insights can inform the development of AI/ML-based Software as a Medical Device (SaMD) for facial phenotyping and may also enhance diagnostic accuracy and clinical utility in future healthcare applications.

To examine how medical geneticists, particularly those with expertise in facial dysmorphology, utilize and are influenced by AI-generated prediction probability and saliency-based XAI in diagnostic decision-making, we solicited board-certified or board-eligible clinicians specializing in medical genetics; a total of 23 and 21 participants who completed the AI-only and XAI-supported diagnosis task, respectively. Specifically, we assessed whether AI-only and saliency-based XAI improved participant performance and confidence by exploring key factors such as the accuracy of AI model classifications and the recognizability of clinical features. Beyond accuracy and confidence improvement, we evaluated how AI and XAI interventions affect geneticist likelihood to follow AI recommendation based on their confidence.

## MATERIALS AND METHODS

### Data collection and image selection

Following our previous work, we used publicly available facial images of individuals with 22q11.2 deletion syndrome (22qDS) (OMIM# 611867), Angelman syndrome (AS) (OMIM# 105830), Kabuki syndrome (KS) (OMIM# 147920, 300867), Noonan syndrome (NS) (OMIM# 163950, 605275, 609942, 610733, 611553, 613706, 615355, 616559, 616564, 619087), and Williams syndrome (WS) (OMIM# 194050), as well as images of unaffected individuals. These conditions were selected due to their recognizable facial features, the variable degree of difficulty in identification based on facial features (e.g., the features of NS may be more recognizable than those of 22qDS), and because they are common enough for most medical geneticists to have first-hand experience with affected individuals.

A total of 3547 images were included in the full dataset: 22qDS (n=591), AS (n=456), NS (n=329), KS (n=247), WS (n=529), unaffected individuals (n=228), and other genetic syndromes (n=1167). Available metadata included age, sex, and ancestry. Eighteen images (three per syndrome and three unaffected) were selected for the user study. These images represented pediatric individuals (newborn to 18 years), varying ancestries, and both sexes. Three clinical geneticists independently reviewed the images to confirm that they accurately depicted one of the five syndromes or an unaffected individual (that is, they would be considered to show a relatively typical presentation of the conditions).

For the study, we selected eighteen images that were unseen during the training of the AI model used to acquire predictions and XAI explanations. These images included two correctly classified and one incorrectly classified image per syndrome, allowing us to approximate classifier performance and validate the effect of AI-only and XAI interventions on both correctly and incorrectly classified samples.

### Models

We trained a ResNet-50 classifier to provide prediction probability for each of six possible label choices: the five genetic conditions and “unaffected”. The label with the highest prediction probability was considered the model’s classification. [11] DeepLIFT,[13] was used to visually highlight areas-of-interest with respect to the predicted label (i.e., saliency maps). A more detailed evaluation of different saliency map algorithms is available in our previous work.[11] The contributions of these areas were numerically summarized as a region relevance score histogram (Figure 1). Being a simplified version of the saliency map, this histogram concisely prioritizes the syndromic areas-of-interest.

**Figure. 1:**
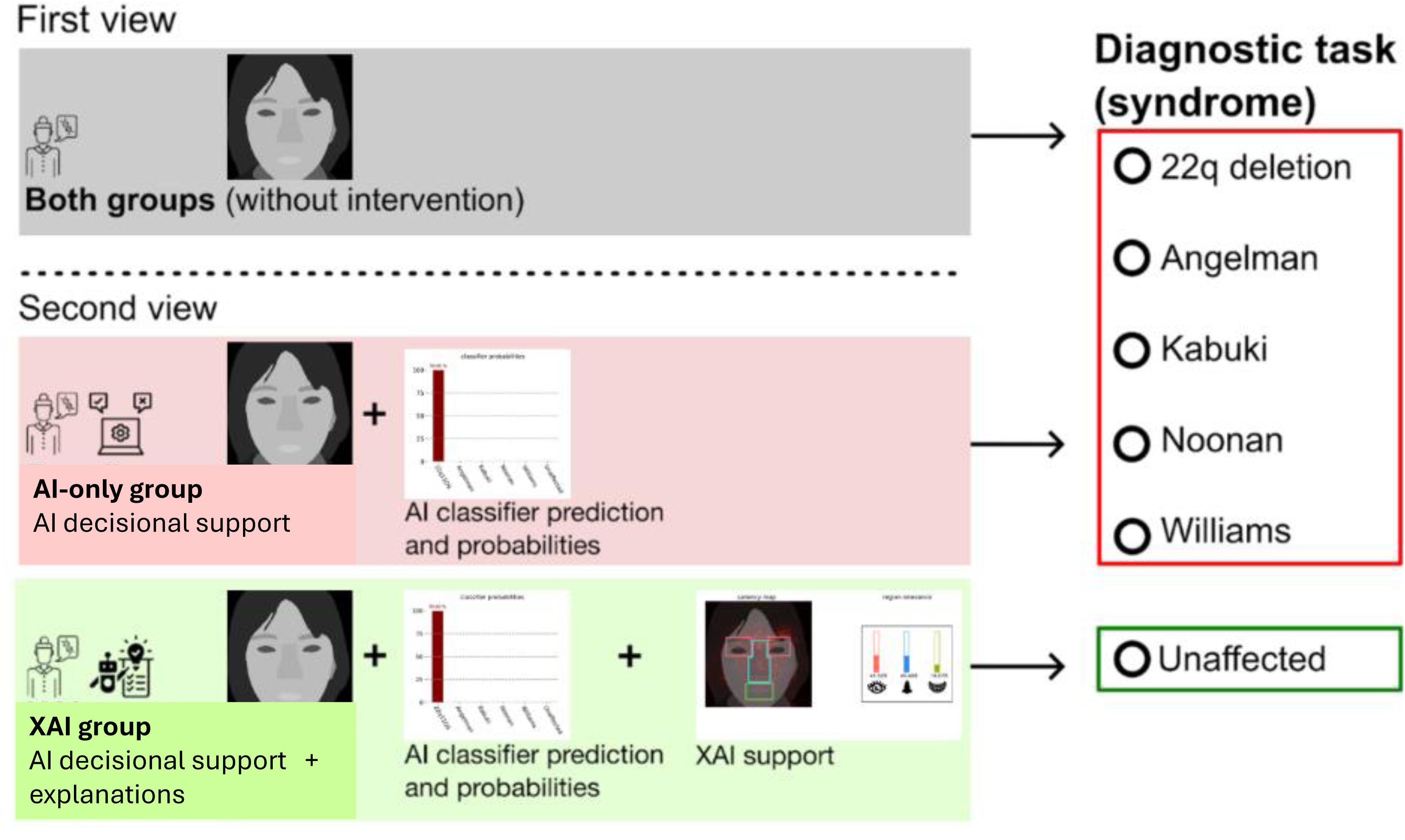
Overview of the AI- and XAI-assisted diagnostic task workflow. In the first view, participants from both groups examined facial images without assistance and selected a diagnosis from five genetic conditions or “unaffected”, while also rating their confidence. In the second view, participants received AI support: the AI-only group saw AI probability, while the XAI group additionally received two explainability features: saliency maps and region relevance scores. After, both groups repeated the diagnostic task, reassessed their confidence, and rated the usefulness of the provided support.

### Recruitment of Participants

Medical geneticists, particularly those with expertise in facial dysmorphology, were recruited for this study (n=126). Participants were identified through professional networks and publications. Recruitment was conducted via email, which provided a link to one of two surveys corresponding to the AI-only or XAI intervention (see descriptions below). Participants were all board-certified or board-eligible physicians who self-identified as medical geneticists or who were in their final month of medical genetics residency/fellowship. See Supplementary Figure 1 and Supplementary Table 1 for participants’ years of experience in genetics and distribution of institutional affiliations. The survey was conducted from June 2024 to July 2025. This study was approved as IRB-exempt by the NIH IRB (IRB# 002178).

### Comparison of survey interventions

We only included results from surveys with at least 9 of the 18 questions completed. This resulted in 44 valid surveys (41 completed all 18 questions). All responses from each participant are treated with equal weight, even if a participant did not complete all 18 questions.

We compared the 2 types of interventions via surveys sent using Qualtrics (Provo, Utah, United States). Surveys were specific to 1 of 2 survey interventions: 1) test images + prediction probability only output (AI-only), 2) test images + prediction probability + XAI tool outputs (XAI). Each survey included 3 images of different individuals with the 5 syndromes and unaffected for a total of 18 images for the participant to classify. Participants were not told prior to the survey the type of intervention they would receive nor the numbers of images of each type.

Both survey versions included a schematic of how AI-based classification generates a prediction probability (percent likelihood an individual has a particular genetic syndrome) and a description of each of the 5 genetic syndromes including a reference image. The XAI version of the survey also included basic information about saliency maps and region relevance, as well as how to interpret these outputs. Participants were shown each of the 18 images one at a time, initially without any intervention and asked to classify the image, as well as rate their confidence level for each classification. They were then shown the image a second time with the classifier’s prediction probability with or without the XAI outputs and asked again to classify the image and rate their confidence level, rate the usefulness of each AI/XAI outputs shown, as well as provide comments (optional). Confidence and rating of usefulness were captured via a 5-point Likert scale (−2 to +2). Participants answered demographic questions (e.g., years of practice) and questions about their use and opinions of AI diagnostic tools. Surveys can be found at: https://github.com/rlwaikel/FacialXAI.

### Evaluation metrics and statistical analysis

Participants in both the AI-only and XAI groups initially viewed the facial images and completed a diagnostic task (selecting the most likely genetic syndrome or the “unaffected’) without AI or XAI assistance and subsequently rated their confidence on a five-point Likert scale. In the second viewing, each group reviewed the same images accompanied by AI predict probabilities with the addition of the XAI tools only for the XAI group. The participants then repeated the diagnostic task and provided confidence ratings. Finally, they assessed the AI tool’s helpfulness across three aspects (prediction probability, saliency maps, and region relevance representations) using a five-point Likert scale. Due to a survey recording error, only 17 of the 18 image results are included in the analyses. To compare accuracy and confidence, we first evaluated the assumptions of normality (Shapiro–Wilk test) and homogeneity of variances (Levene’s test) to determine the appropriate statistical test. When both assumptions were satisfied, we used the parametric T-test. If either assumption was violated, we instead used the non-parametric Mann–Whitney U test. Mediation analyses were further performed to analyze the direct and indirect effects of the AI predicted label probability, saliency maps, and relevance scores on the user decision to follow AI. A detailed explanation of experimental design is provided in Supplementary Methods.

## RESULTS

### Impact of AI- vs. XAI-assisted decision-making diagnostic performance and confidence conditioned on model accuracy

To compare the impact of AI and XAI on accuracy, we first divided images according to whether the classifier’s classification was correct (true) or incorrect (false), as the ground truth could impact participant’s behavior (Figure 2A). When the AI model classification was correct, accuracy improvement in selecting the correct genetic condition or unaffected was similar between both groups (0.20± 0.13 (AI-only) and 0.19±0.13 (XAI)). Conversely, when the AI model classification was incorrect, there was a statistically significant decrease in accuracy as compared to correct AI predictions for both the AI-only (−0.20±0.22 vs. 0.20± 0.13; U=15.0, p<0.0001) and XAI group (−0.21±0.23 vs. 0.19±0.13; p<0.0001). Additionally, there was a statistically significant increase in reported confidence by participants for images the model classified correctly vs. incorrectly after exposure to both the AI-only (0.30±0.26 vs. −0.14±0.64, p=5.1^-3^) and XAI (0.38±0.71 vs. −0.14±0.76; U=113.5, p=7.2×10^-3^) decisional support tools (Figure 2B).

**Figure 2:**
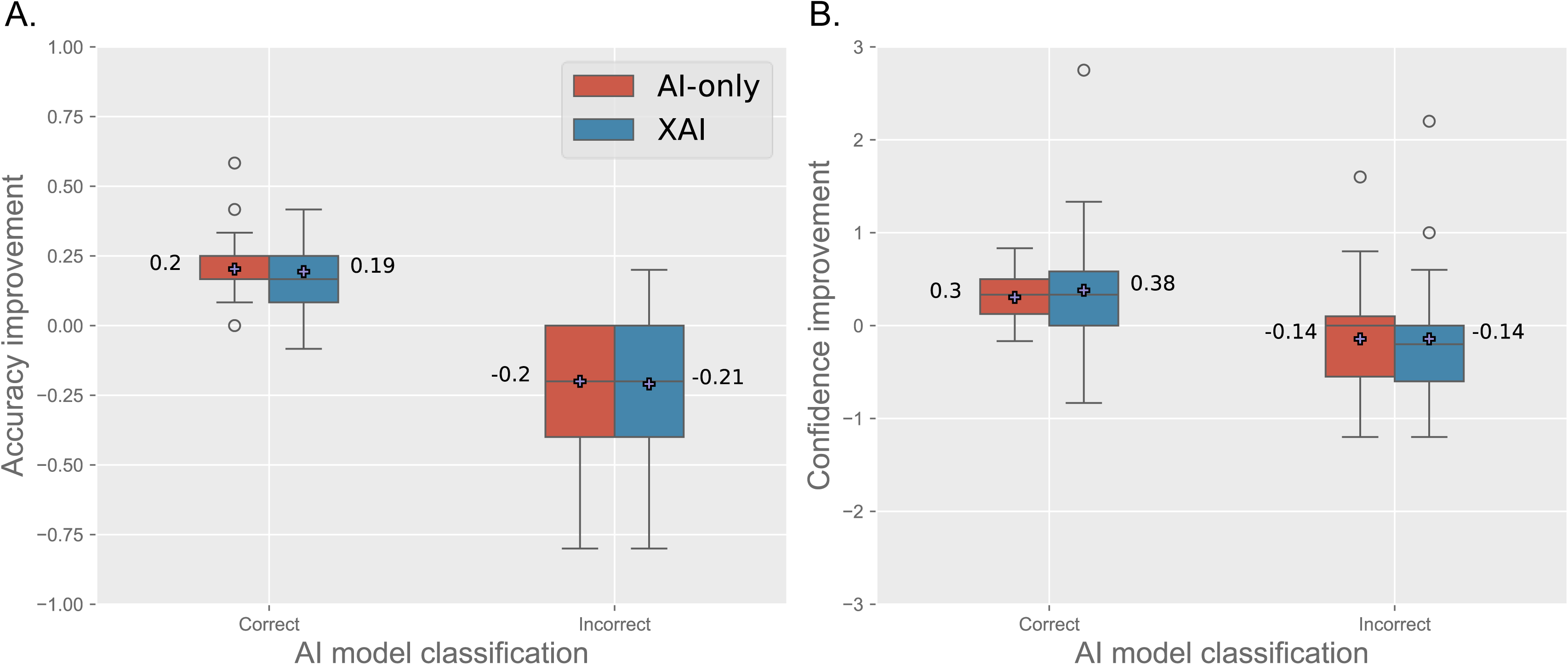
AI model classification accuracy affects expert user’s accuracy and confidence. Accuracy improvement (left) and confidence improvement (right) are divided between when the AI model’s classification were correct and incorrect. In both AI-only and XAI groups, interventions positively impacted participants’ accuracy when the AI model was correct. However, when the AI model made incorrect classifications, participants’ performance and confidence decreased.

### Impact of AI- vs. XAI-assisted decision-making diagnostic performance and confidence conditioned on initial participant performance

Participants’ responses may be affected by whether an image shows more obvious dysmorphic facial features and thus would be easier to diagnose. Assuming facial images of individuals with more obvious dysmorphic facial features would have higher participant accuracy, we divided images according to the initial accuracy of all participants prior to seeing AI or XAI decision help intervention. Images for which > 70% of participants selected the correct genetic conditions were designated as “obvious” dysmorphic features (n=9) and the rest, which had <62% accuracy (n=8) were designated more “difficult” (Figure 3). Average accuracy improvement did not statistically differ for both obvious images and more difficult images in both the AI-only (0.07±0.11 vs. 0.10±0.16; U=214.5, p=0.27) and XAI interventions (0.04±0.14 vs. 0.12±0.19; U=149.0, p=0.06).

**Figure 3:**
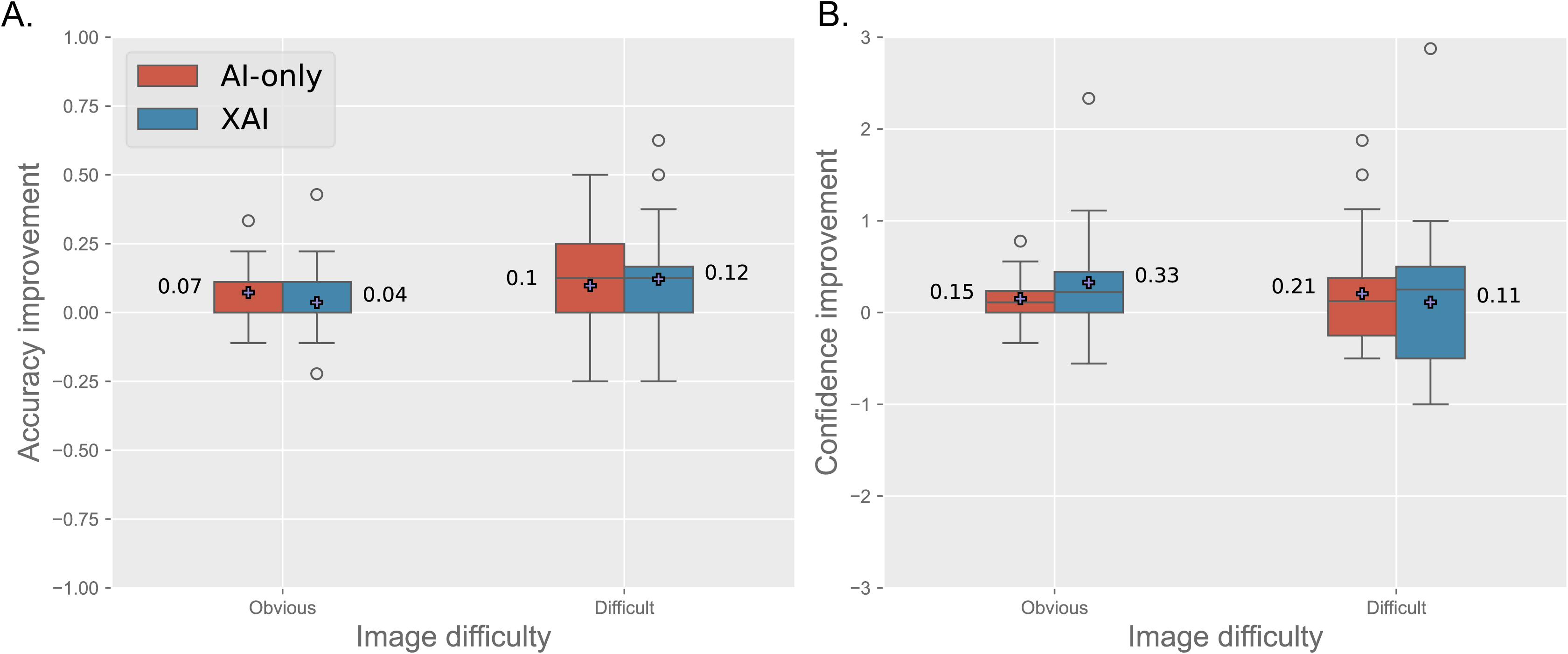
User accuracy and confidence improvement according to difficulty of image diagnosis. Accuracy improvement (left) and confidence improvement (right) are grouped by cases where more obvious clinical features are present as defined by greater than 70% of participants were correct in their diagnosis without AI assistance: easier to decern for clinicians (9 images) versus more challenging samples with increased difficulty (8 images) as defined by less than 62% of participants were correct without AI assistance. All values are aggregated per participant.

Although there is an increase with the confidence score, due to large standard deviations, we did not reach a statistical significance for XAI-group (0.33±0.59 vs. 0.11 ±0.84, U=253.0, p=0.42) nor AI-only group (0.15±0.24 vs 0.21 ±0.60, U=273.0, p=0.86).

(Figure 3B). Of note, 5 of the 6 AI-misclassified images were in the participants’ more difficult group.

### Perceived usefulness of AI model output and explanations

Overall, participants rated AI predicted probabilities as useful, regardless of whether these participants picked the correct answers, or whether predictions were accurate (see the generally positive trend in the Figure 4A histogram and Supplementary Table 2A). The usefulness scores of seeing the AI predictions among the AI-only and XAI participants on average were 0.60±1.1, and 0.74±1.3, respectively. The reverse trend is seen with saliency maps and region relevance score usefulness (Figure 4B). On average, the XAI responses yielded negative scores for these two metrics −0.12±1.4 and −0.17±1.3, respectively. We note that the majority of participants either viewed XAI as unhelpful or had a neutral perception. Approximately one-quarter of participants (25% and 27%) selected a −2 score (the lowest option) for saliency maps and region relevance scores, respectively (Figure 4B). This finding is also reflected in the open-ended user feedback in Supplementary Table 2A.

**Figure 4:**
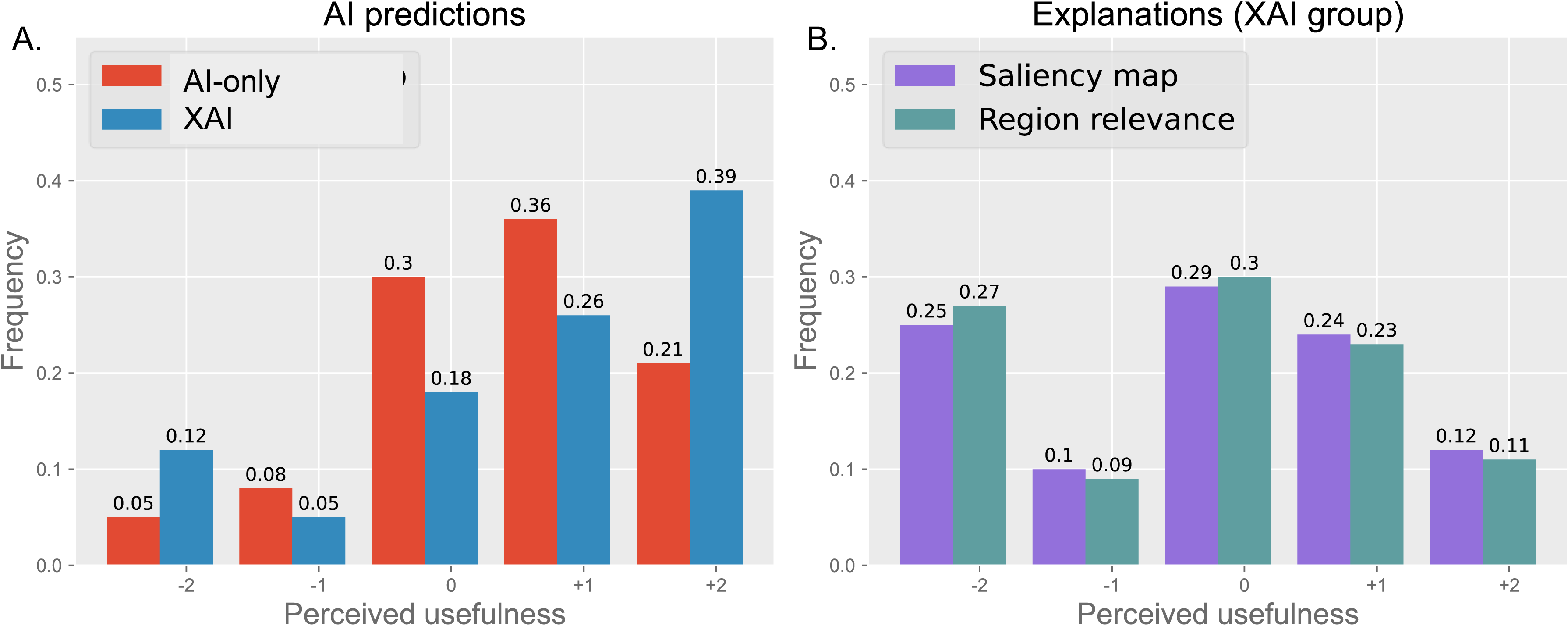
Perceived usefulness of AI and XAI outputs. Participants’ perceived usefulness of AI prediction probability (left), saliency maps, and region relevance scores(right). Overall, respondents in both the AI-only and XAI groups found AI predictions more useful than saliency maps and region relevance scores. In the XAI group, the ratings of saliency maps and region relevance scores were nearly identical.

For AI-only and XAI cohort, divided between AI-correct (Table 1A) and AI-incorrect (Table 1B) image sets, we computed the pairwise correlations for these four metrics: accuracy improvement, prediction probability usefulness, saliency map usefulness, and region relevance score usefulness. In both the AI-incorrect and AI-correct image set and for both the AI-only and the XAI cohort, we found significant positive pair-wise correlations between the usefulness scores of prediction probability, saliency map, and region relevance score (All p-values <0.001, for the Spearmans’ Rho values see Table 1). For the AI-incorrect image set, we found significant negative correlations between accuracy improvement and probability usefulness (AI-only, rho= −0.31, p=0.001; and XAI, rho= −0.40, p<0.001).

**Table 1A:** Correlation analysis between participants’ perceived usefulness ratings and accuracy improvement in AI-correct subset. Spearman’s Rho, degrees of freedom, and p-values are reported.

| *AI-only group* |  | Prediction<br>usefulness | Saliency<br>usefulness | Region<br>usefulness |
| --- | --- | --- | --- | --- |
| **Accuracy**<br>**improvement** | **Spearman's Rho** | **0.11** | **—** | **—** |
|  | **df** | **264** | **—** | **—** |
|  | **p-value** | **0.066** | **—** | **—** |
| *XAI Group* |  |  |  |  |
| **Accuracy**<br>**improvement** | **Spearman's Rho** | **-0.032** | **0.098** | **0.096** |
|  | **df** | **250** | **250** | **250** |
|  | **p-value** | **0.609** | **0.122** | **0.132** |
| **Saliency**<br>**usefulness** | **Spearman's Rho** | **0.37** | **—** | **—** |
|  | **df** | **250** |  |  |
|  | **p-value** | **<.001** |  |  |
| **Region**<br>**usefulness** | **Spearman's Rho** | **0.45** | **0.87** | **—** |
|  | **df** | **250** | **250** |  |
|  | **p-value** | **<.001** | **<.001** |  |

**Table 1B:** Correlation analysis between participants’ perceived usefulness ratings and accuracy improvement in AI-incorrect subset. Spearman’s Rho, degrees of freedom, and p-values are reported.

| *AI-only group* |  | Prediction<br>usefulness | Saliency<br>usefulness | Region<br>usefulness |
| --- | --- | --- | --- | --- |
| Prediction<br>improvement | Spearman's Rho | -0.31 | — | — |
|  | df | 109 | — | — |
|  | p-value | 0.001 | — | — |
| *XAI Group* |  |  |  |  |
| *Accuracy*<br>*improvement* | Spearman's Rho | -0.40 | -0.015 | -0.078 |
|  | df | 103 | 103 | 103 |
|  | p-value | <.001 | 0.881 | 0.431 |
| *Saliency usefulness* | Spearman's Rho | 0.43 | — | — |
|  | df | 103 |  |  |
|  | p-value | <.001 |  |  |
| *Region*<br>*usefulness* | Spearman's Rho | 0.48 | 0.91 | — |
|  | df | 103 | 103 |  |
|  | p-value | <.001 | <.001 |  |

Furthermore, with the AI-only cohort, when the AI classification was correct, then there was a mediated effect by the AI predicted label probability from user initial confidence toward their decision to Follow AI. The XAI components (saliency maps and relevance scores) did not have significant effects on the user decision to Follow AI (See Supplementary Results).

### Users’ Qualitative Feedback on AI and XAI Support

In addition to determining whether there was a quantitative difference between AI and XAI support, we also solicited qualitative opinions about these interventions. At the end of the survey, we included an optional open-ended question asking each participant to provide feedback on the support tools they used during the survey. Many participants included feedback (15/23 and 19/21 for the AI-only and XAI groups, respectively) (see Supplementary Table 2A and respective word cloud representations in Supplementary Figure 5).

For this question, we asked participants to consider how much they relied on the tools when making their decision. To quantify the results, key words and phrases (e.g., helpful/not helpful) were manually identified to determine whether participants demonstrated a positive view of AI tools in their decision making. Of the 15 participants in the AI-only group, 66.7% (n=10) reported positive opinions of the prediction probability in their decision-making, whereas 20% (n=3) had negative opinions, and 13.3% (n=2) provided neutral opinions. In contrast, of the 19 participants in the XAI group, only 42.1% (n= 8) reported positive opinions (primarily towards the AI prediction probability), 47.4% (n=9) had negative opinions, and 10.5% (n=2) had neutral opinions. While a description of how to use XAI was included in the introductory part of the survey, 3 XAI survey participants reported not knowing how to use the tools.

## DISCUSSION

### Impact of AI and XAI support on diagnostic performance

#### AI improved diagnostic performance when it was correct

With respect to accuracy, similar trends were seen for both AI-only and XAI participants. When AI predictions were accurate, participants were more likely to change their initial incorrect answer to the correct answer in the second viewing (Figure 2). In contrast, false AI predictions either had no significant effect or misled participants into selecting an incorrect answer in the second viewing. Potentially, if a participant initially selected an accurate answer but lacked familiarity with the conditions, then this participant might have been misled by an incorrect AI classification in the second viewing. In such a situation, XAI could be valuable since users can inspect the areas-of-interests determined by the AI classifier. However, this was not the case as explained below and in the section “*XAI did not significantly enhance user performance and perception”.* However, the standard errors are large, and more studies (e.g., with more participants) would be needed to assess this hypothesis.

One possible reason for AI’s negative impact on incorrectly classified cases is the model prediction probability. For example, an image showing a person with KS was incorrectly predicted to be 22qDS with a prediction probability of 89.5% (Figure 5B). This overconfidence may mislead users, reinforcing incorrect diagnoses.

**Figure 5:**
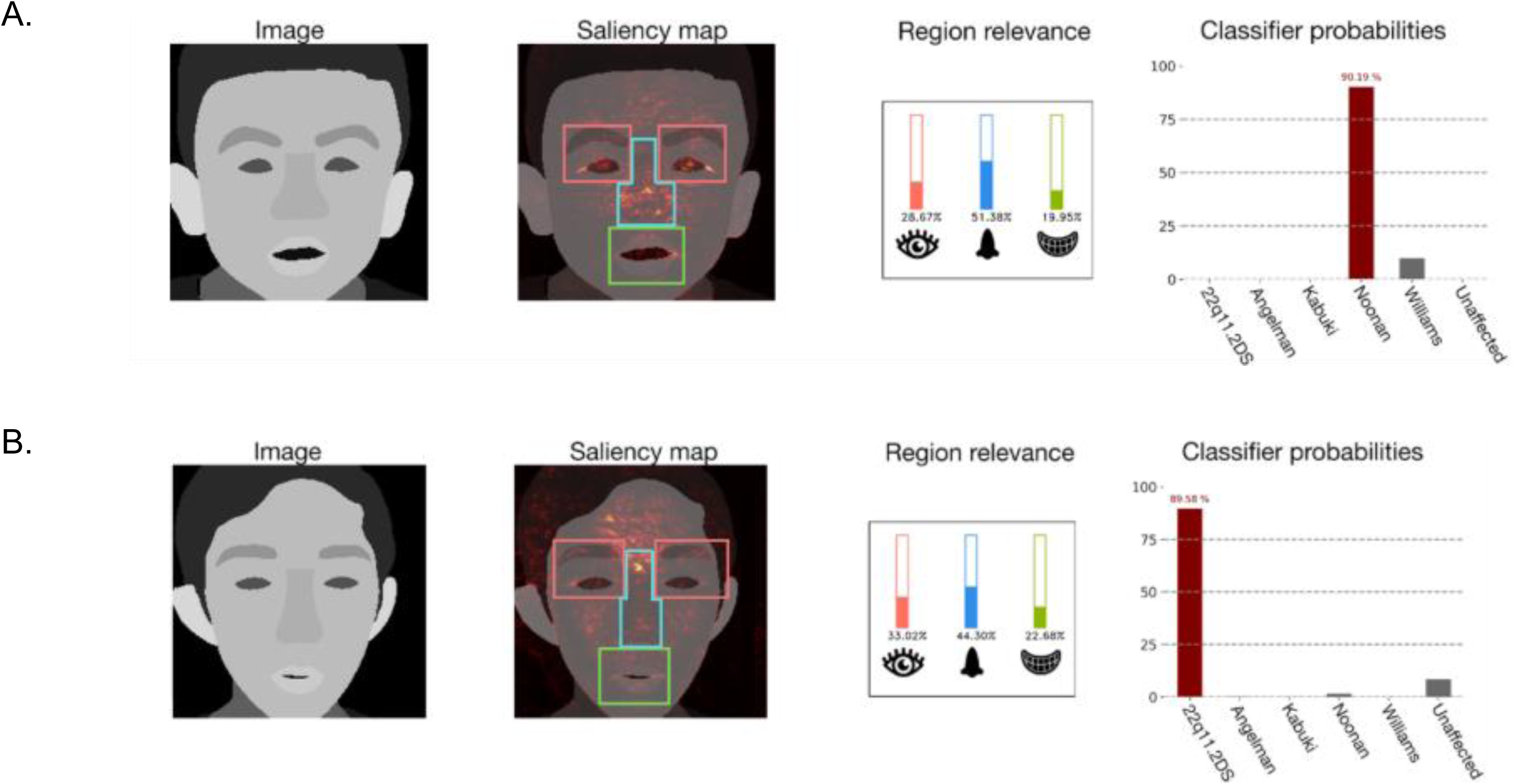
Examples of AI model prediction probability, saliency maps and region relevance score histograms. An example of the AI model’s accurate classification of an individual with Noonan syndrome is shown in panel A. An example of the AI model’s inaccurate classification of an individual with KS; incorrectly classified as 22qDS is shown in B.

#### XAI did not significantly enhance user performance and perception

On average, the XAI intervention method in our study did not lead to a performance improvement compared to the AI-only cohort. One potential reason for this is that participants were always required to decide in our study. In real practice, they would likely seek additional information about the patient when they are unconvinced by the explanations. The low user satisfaction with XAI and the bigger decrease in median confidence for XAI in Figure 2B indicates that the explanations made the participants more critical towards incorrect AI-predictions. However, they could not act on this suspicion to get more information. While carefully-controlled experiments can be efficient and cost-effective, these observations highlight that caution is required when attempting to extrapolate results to real-world settings.[14]

Nevertheless, our findings are consistent with previous studies, which also did not show significant improvements from saliency maps.[7 15] Similar to our low perceived usefulness ratings and the negative textual feedback regarding saliency maps, those studies also found that saliency maps were difficult to interpret and increased users’ cognitive load. Other XAI-assisted strategies may be more intuitive for users. For instance, Mertes et al. found counterfactual explanations to be significantly more useful than saliency maps in a pneumonia classification task,[7] However, their task is arguably simpler than identifying rare genetic conditions. Therefore, future studies would be needed to confirm whether counterfactuals are helpful in the context of rare genetic conditions. There may be other nuances entirely unrelated to the XAI methods. For example, we previously found that expert clinicians displayed unique eye movements when analyzing the same facial images.[16] In a few instances, the clinicians could still identify the correct disease although their eye movements differ from each other and from the model saliency maps.

#### Participants did not differentiate saliency maps and region relevance scores

The region relevance score histogram summarizes the facial areas-of-interest contributions toward the model prediction. Hence, when compared against a saliency map, the region relevance score histogram is a simpler interpretation about how the model works. However, the participants did not prefer the region relevance scores over saliency maps, or vice versa (see correlations in Table 1).

#### Insights from example images

To gain deeper insights into the potential benefits and risks of XAI approaches, we examined two images that showed a strong difference in accuracy improvement between the AI-only and XAI groups (Supplementary Figures 2 and 3). Members of our group with expertise in clinical dysmorphology provided their interpretation of these examples.

Figure 5A shows an example of an individual with NS where the AI made a correct classification of NS. In this case, the XAI group exhibited both greater accuracy improvement and higher confidence in their prediction. One expert commented that the down slanting palpebral fissures, a common finding in NS, were highlighted in the saliency map (see Expert Commentary 1 in Supplementary Table 2B). This suggests that the saliency map aligns well with the AI’s prediction, which may explain why the explanation increased participants’ confidence in the AI’s correct decision.

Figure 5B shows an individual with KS where the model incorrectly predicted to be 22qDS. In this case, both the AI and XAI groups showed negative accuracy improvement, but the decline was less pronounced in the AI-only group. The confidence in this wrong classification was lower in the XAI group. One expert noted that this KS image can easily be misclassified as 22q11DS (see Expert Commentary 2 in Supplementary Table 2B). This image’s saliency map appears reasonable with respect to 22qDS because of the highlighted nasal bridge typically seen in 22q11DS. While the saliency maps also highlighted KS features, such as areas around the eyes,[17] these might not be well known among clinicians according to this expert. While anecdotal, this example suggests that domain experts may provide useful insights in designing and evaluating clinical XAI studies.

#### Mediation Analysis on effects of XAI Presence

We found that XAI Presence does not directly or indirectly influence participants’ decisions to Follow AI. Rather, the users rely on initial intuitions (e.g., their initial confidence level) when deciding to follow AI for both AI-correct and -incorrect image subsets.

### Limitations of our study

This study explored the applicability of XAI in clinical genetics but acknowledge limitations.

First, the cohort size, type, and inherent biases could impact results. We had a small sample size in part due to the limited number of dysmorphology experts, as well as low participation rates, likely due to clinicians’ busy schedules. Clinicians’ prior knowledge and familiarity with XAI could have influenced their perceptions. There may also be human-specific patterns to assessing faces; XAI studies involving faces may be affected by these patterns. As a future study, it would be interesting to compare results across different image types used in medical genetics (e.g., faces, skeletal X-rays, and skin findings).[18–20]

Second, DeepLIFT saliency maps only visually highlight areas-of-interest which, in some instances, can appear unrelated to the three general syndromic face regions (eyes, nose, and mouth). Our region relevance score method succinctly and numerically assessed these three specific facial areas. There are many relevant detailed phenotypes (i.e., our previous study, [11] indicated 50 phenotypes associated with these genetic conditions). Pointing out specific important features (such as based on Human Phenotype Ontology terms) could have better helped users identify the genetic conditions. For example, future studies such as vision-language models,[21] may make explanations more understandable. Also, future studies could investigate alternative XAI methods, like counterfactual explanations,[7] and concept bottleneck models[22].

Lastly, our classifier is based on ResNet, and the choice of model architecture can influence saliency maps. Due to our small dataset, we opted for ResNet instead of other larger networks such as Vision Transformer.[11] Future studies could include exploring other DL architectures with larger datasets. Beside the model architecture, saliency map effectiveness may also depend on the model’s performance; for example, when the model misclassified an image, the saliency map with respect to this ground-truth label may not accurately highlight the syndromic features. In this case, one should not trust the saliency map.[10 23] However, ground-truth labels are often unavailable when recognizing syndromic faces in a clinical setting. Hence, in practice, we cannot anticipate whether model classification is accurate; thus, inspecting the saliency maps is sensible.

## CONCLUSIONS

Our main objective was to establish a framework for assessing the impact of explanations on decision-making, rather than to evaluate specific XAI techniques. While our AI/XAI support was perceived as useful, especially in cases where it aligned with participants’ diagnostic reasoning, it had limited impact on overall diagnostic performance. Recent work has underscored the heterogeneous outcomes of human-AI collaboration across tasks and domains, with some scenarios showing a performance decline when AI is introduced.[24] For facial phenotyping, which remains central in clinical genetics, our study suggests a potential for synergy, indicating that AI explanations can support clinicians when appropriately aligned with their diagnostic process. To address this potential, future research should address sources of confusion and skepticism identified by users and conduct larger-scale studies to understand the scenarios in which XAI enhances diagnostic accuracy.

## Supporting information

NIH Publishing Agreement

## Data Availability

Code to train model and make saliency maps are at https://github.com/sumeromer/facial-gestalt-xai. Code to reproduce figures and statistical tests for surveys are at https://github.com/rlwaikel/FacialXAI. Raw results of the user survey responses are also available on the project repository.

https://github.com/sumeromer/facial-gestalt-xai

## ACKNOWLEDGEMENTS

This research was supported by the Intramural Research Program of the National Human Genome Research Institute, National Institutes of Health and Discovery Grant (2022-03727) of Natural Sciences and Engineering Research Council of Canada (NSERC). It has also been partially funded by the Deutsche Forschungsgemeinschaft (DFG) through the Leibniz Award of Elisabeth André (AN 559/10-1). The contributions of the NIH author(s) were made as part of their official duties as NIH federal employees, are in compliance with agency policy requirements, and are considered Works of the United States Government. However, the findings and conclusions presented in this paper are those of the author(s) and do not necessarily reflect the views of the NIH or the U.S. Department of Health and Human Services. This research used the computational resources of the National Institutes of Health High-Performance Computing Services Biowulf cluster.

## AUTHOR CONTRIBUTIONS

O.S. contributed to conceptualization, data curation, formal analysis, investigation, methodology development, project administration, and validation. T.H. was involved in data curation, formal analysis, methodology, validation, and drafting the original manuscript. J.C. participated in methodology, formal analysis, validation, and reviewing and editing the manuscript. D.D. participated in data curation, methodology development, software development, validation, and reviewing and editing the manuscript. S.L.H. contributed to resource provision and manuscript review and editing. C.C. was involved in conceptualization, methodology development, funding acquisition, supervision, and reviewing and editing the manuscript. E.A. contributed to conceptualization, methodology development, funding acquisition, supervision, and manuscript review and editing. B.D.S participated in funding acquisition, methodology development, resource provision, supervision, and reviewing and editing the manuscript. R.L.W. contributed to conceptualization, data curation, investigation, methodology development, project administration, resource provision, supervision, and both original drafting and editing of the manuscript.

## DATA AND CODE AVAILABILITY

Code to train model and make saliency maps are at https://github.com/rlwaikel/FacialXAI. Code to reproduce figures and statistical tests for surveys are at https://github.com/rlwaikel/FacialXAI. Raw results of the user survey responses are also available on the project repository. Code for mediation analysis is https://github.com/jacie-cheng/FacialXAI_Mediation_Analysis. OS has full access to all of the data in the study and takes responsibility for the integrity of the data and the accuracy of the data analysis.

