## Supplemental Text for "Application of deep learning and explainable AI-supported medical decision-making for facial phenotyping in genetic syndromes"

**Supplementary Methods**

This experimental design enabled a comparison of three key measures before and after the intervention in both groups:

- Performance improvement was assessed by comparing diagnostic accuracy between the first and second views, with possible outcomes of -1 (True→False), 0 (True→True or False→False), or +1 (False→True). In other words, if participants change their previously correct answer to a wrong choice, then the outcome was -1. Conversely, the outcome would be +1 if participants changed their previously incorrect answer to the correct answer. When participants' answers for the two views stayed either correct or incorrect (whether or not their initial choice was correct or incorrect), then the outcome score was 0.
- Confidence improvement was quantified as the difference in self-reported confidence scores, ranging from -2 (“not confident”) to +2 (“highly confident”), between the first and second views (i.e., the view without XAI guidance vs. the one with XAI guidance).
- Usefulness of AI and XAI support was only rated after the second viewing on a Likert scale ranging from -2 (“not useful”) to +2 (“very useful”). The AI-only group evaluated only the helpfulness of seeing just the prediction probability. In contrast, the XAI group assessed the usefulness of seeing both the prediction probability and the saliency map and region relevance explanations.

We compared accuracy and confidence improvements between AI-only and XAI groups before and after the intervention, reporting mean values and confidence intervals (1.96 ~~1.06~~ times the unbiased standard error of the mean). These measures were analyzed by aggregating participant-level data, particularly in two situations: (i) when AI model predictions were true or false, and (ii) across the groups of images containing more obvious clinical features and more challenging images, where dysmorphic features were more difficult to discern. Statistical testing was conducted only to compare these measures. For hypothesis testing between the AI-only and XAI groups, such as differences in accuracy improvement, we first evaluated normality and equality of variance using the Shapiro-Wilk and Levene's tests. Based on these results, either an independent t-test or a Mann-Whitney U-test was applied as appropriate.

Since the AI-only group did not have usefulness ratings for explanations, we did not compare these ratings between groups. Instead, we conducted a correlation analysis to examine the relationship between participants' perceived usefulness of each component and accuracy improvement. Given that both variables were ordinal and do not follow a normal distribution, we used Spearman's correlation, which assesses whether greater accuracy improvement was associated with higher perceived usefulness, even in the absence of a strictly linear relationship. This analysis relies on two key assumptions: (1) each test image was approximately independent and exchangeable, and (2) participants have comparable experience with genetic conditions. These assumptions allow us to relate the aggregated usefulness ratings and accuracy improvement ratios within each group.

Various factors may influence participants' confidence and trust in AI and XAI interventions beyond those captured by diagnostic tasks, confidence ratings, and usefulness assessments. To explore these additional factors, we collected open-ended feedback at the end of the survey. A qualitative analysis of these responses highlighted potential strengths and limitations of AI-assisted facial phenotyping, providing insights to guide future research.

**Supplementary Text – Mediation Analysis**

*Mediation analysis on following AI*

The main analysis did not identify a significant difference in accuracy between or within the AI and XAI cohorts. Hence, in this new analysis, we were no longer concerned with participant accuracy. Rather, we evaluate how the average participant follows AI predictions in different circumstances. We define the term “Follow AI” as below.

Each participant sees the test images twice. First, they see the image alone and then provide their Prior Answer (their most likely diagnosis) and their Prior Confidence in that answer. Second, they see the image and the AI predicted label and then provide their Subsequent Answer and their Subsequent Confidence in that answer. Follow AI was computed based on the participant’s Prior and Subsequent Answers and the AI predicted label. For example, suppose the participant’s Prior Answer does not match the AI predicted label, then the participant’s original choice differs from the AI label. Next, suppose the participant’s Subsequent Answer matches the AI label, then presumably the participant must have followed the AI recommendation (in this case, the Follow AI node in Supplementary Figure 6 was defined as “yes”). However, there were ambiguous situations where we cannot be sure whether the participant follows AI (Supplementary Table 3). Thus, for Follow AI, we exclude these ambiguous cases. This reduced the number of total responses from 726 to 315.

As in the main text, we further divide the entire data according to which images were correctly predicted by the AI classifier. Thus, there were 4 subsets: XAI cohort with AI-correct image subset, and XAI cohort with AI-incorrect image subset (and likewise for the AI-only subset). When the context was clear, for brevity, we will denote the subset of XAI cohort with AI-correct image subset as XAI AI-correct. Similarly, there are also XAI AI-incorrect, AI-only AI-correct, and AI-only AI-incorrect subsets.

On each subset, mediation analysis was done to understand how an average participant may Follow AI (as defined above).[1] Although the mediation analyses were done separately on each subset, we can still compare the results for the subsets. The main objective in this part of the study was to evaluate the direct, indirect, and total effects of each path in the diagram (Supplementary Figure 6).[2, 3] Mediated variables were fitted with linear regressions, and the outcome Follow AI was fitted with logistic regression (Supplementary Figure 6). Our mediation analysis implementation is available at: https://github.com/jacie-cheng/FacialXAI_Mediation_Analysis. Our causal mediation analysis assumes that, within each subset, the images were independent and interchangeable.

Supplementary Tables 4-6 show the regression coefficients from the mediation analyses. In mediation analysis, the indirect effect of exposure X via mediator M to outcome Y was computed as a product of the coefficients $\beta_{XM}$and $\beta_{MY}$[1]. In this analysis, it was necessary to evaluate the statistical significance of this entire product $\beta_{XM}$and $\beta_{MY}.$[4] Thus, bootstrap sampling was done to compute the standard deviations of every indirect as well as the total effect.[5]

The resampling was performed with respect to each image. For each image, we resampled the observations with replacement from its original dataset. Thus, each bootstrap sample contains the same number of observations per image as the original observed dataset. After 1000 iterations, bootstrap samples with outlier regression coefficients (defined as those with 1.5 times their respective interquartile ranges) were removed. The standard deviations of the indirect and direct effects were then computed with the remaining bootstrap samples.

With mediation analysis, for example, in the XAI group, we aim to approximate whether participants follow AI recommendations because they feel more confident after XAI exposure (Supplementary Figure 6). In Supplementary Figure 6, the only source node was Prior Confidence (e.g., all the paths can be traced back to this node). Presumably, user Prior Confidence was the only major factor influencing Follow AI. For example, highly confident participants may not need or trust any XAI component at all. Conversely, low confidence participants may follow AI recommendation because they find the XAI presentation to be informative. Hence, we keep the XAI components as mediators. For brevity, we name the XAI components in the mediation diagram as: Map, Relevance Score, and Probability for the helpfulness of saliency map, relevance score histogram, and seeing AI predicted probability of the disease, respectively.

*Mediation analysis in AI-only cohort*

Conditioned on the AI-only participants, in both AI-correct and -incorrect subsets, participant Prior Confidence has different direct effects on Follow AI. In the AI-correct subset, high Prior Confidence participants were statistically less likely to directly Follow AI, whereas this effect was not significant in AI-incorrect (Supplementary Table 4B Row 1 and Supplementary Table 4D Row 1). The same trend was seen for the indirect effect of the path Prior Confidence → Probability →Follow AI. Here, statistical significance was only seen in AI-correct subset not in AI-incorrect subset (Supplementary Table 4B Row 3 and Supplementary Table 4D Row 3).

Interestingly, the AI-only AI-correct subset displays inconsistent mediation [8], where the direct and indirect effects have opposite signs (e.g.,-1.2 vs 0.58 in Table 4B).  For example, high Prior Confidence participants were less likely to Follow AI, although these participants tend to not rate Probability very negatively (e.g., helpfulness scores can still be above zero). Conversely, low Prior Confidence participants were more likely to Follow AI although these participants tend to not rate Probability very positively (e.g., helpfulness scores can still be below zero). Also, due to inconsistent mediation, the total effect from Prior Confidence to Follow AI was not different from zero (Supplementary Table 4B Row 5).

*Mediation analysis in XAI-cohort*

Conditioned on the XAI-cohort only, between the AI-correct against AI-incorrect subset, we do not see any differences in Prior Confidence → Follow AI or Prior Confidence -→ Probability → Follow AI relationship. There were also no other differences between the two subsets (Supplementary Tables 5B and 5D). None of the direct or indirect mediated paths were statistically significant.

Compared to AI-correct subset, images in AI-incorrect subset were subjectively harder to recognize (based on an assessment of the images by the participating geneticists). Regardless of the image difficulty level, however, when provided with XAI findings, the participants cannot understand how saliency maps and region relevance scores should interact with the model predicted label. Hence, there was no direct association between their Prior Confidence and Follow AI, nor any indirect effects from Prior Confidence via any XAI component toward Follow AI. This result was perhaps unsurprising since many open-ended survey responses about XAI were negative.

*Cross-comparing mediation analysis between XAI and AI cohorts*

There were no differences when cross-comparing AI-only versus XAI in the AI-incorrect image subset (Supplementary Tables 4D and 5D). However, there were differences between the AI-only and XAI cohorts in the AI-correct subset. Unlike the AI-only AI-correct subset, the XAI AI-correct subset does not show a significant direct effect for Prior Confidence → Follow AI, nor a significant indirect effect for Prior Confidence → Probability → Follow AI (Supplementary Table 5B, Row 1 and 5).

We hypothesize an explanation for the differences between the AI-only and XAI cohorts in the AI-correct subset. Images in the AI-correct subset may be easier to diagnose. In this scenario, without XAI components, participants can still assess the model Probability based solely on their Prior Confidence and then decide to Follow AI. However, as noted in the previous section, when exposed to XAI, the participants appear to have difficulty understanding how the saliency maps and relevance scores interact with the model prediction, and thus the participants could not rely on the model Probability as a contributor for their Follow AI decision.

*Mediation analysis on AI-correct user-hard image subset*

When AI classification was incorrect, user behavior can be intractable; for example, in the case of AI hallucinations, some people still blindly trust AI.[6] Following this observation, this section estimates how AI-correct predictions can help the participants in tough situations. Hence, we narrow down the dataset to only 6 (out of 12) AI-correct images. Images 2, 4, 6, 7, 13, and 15 were deemed difficult by the 44 participants based on their average Prior Accuracy (Supplementary Table 7). Because of the limited number of samples in this subset, bootstrap standard deviations were calculated based on 200 bootstrap iterations, instead of 1000. We denote this subset as AI-correct user-hard and conduct mediation analyses following the diagrams in Supplementary Figure 6.

In the AI-only cohort, the direct effect Prior Confidence → Follow AI was negative and significant, indicating that highly confident users were unlikely to follow AI based on their initial intuition, and vice versa. Interestingly, the indirect effect Prior Confidence → Probability → Follow AI was positive and significant. Thus, highly confident participants still tend to Follow AI after being presented with a correct AI prediction. We did not observe any significant effect involving Subsequent Confidence, indicating that users do not Follow AI because they feel more confident afterward (Supplementary Table 8B).

In the XAI-cohort, there were no significant mediation paths. Thus, even when the AI classification was correct, the XAI components did not increase the likelihood of Follow AI. Here, the Probability was also no longer having any significant effect on Follow AI (Supplementary Table 9B).

Because this image subset contains AI-correct user-hard cases, in the AI-cohort, it was not surprising for the model Probability to have an impact on user behavior. However, in the XAI-cohort, the XAI components did not lead to any significant mediation paths, including paths involving model Probability. Arguably, showing XAI might have resulted in an unintended negative “side effect”, perhaps because viewers were confused by or did not value XAI.

*Mediation analysis on XAI Presence and Follow AI*

This section explores the XAI Presence effect on user decision to follow AI. This mediation analysis uses all 44 participants; however, the images were still partitioned into 2 subsets: AI-correct and -incorrect. In particular, each subset would contain responses from 44 participants; the participants will be either exposed to XAI components or not. XAI Presence was a binary variable (e.g., 0 or 1) to define whether the XAI components (i.e., the saliency maps and relevance score histograms) were shown (Supplementary Figure 6C). Thus, this section objectively measures the XAI Presence effect on Follow AI and does not rely on helpfulness scores which can be subjective.

The AI-correct subset has a significant indirect effect on the relationship Prior Confidence → Subsequent Confidence → Follow AI that was not seen in the AI-incorrect subset (Supplementary Tables 6B and 6D, Row 2). Although the effect was very small, the AI-incorrect subset also has a significant negative direct effect Prior Confidence → Follow AI that was also seen in the AI-correct subset (Supplementary Tables 6B and 6D, Row 1). Thus, in both subsets, low confidence participants are still more likely to follow AI regardless of how they feel after seeing the model predicted labels, and vice versa. There were no other significant paths involving XAI Presence in either subset. Thus, XAI Presence does not appear to influence participants’ decisions to follow AI (Supplementary Table 6B and 6D, Rows 4-6).

Since the model Probability was always shown regardless of the XAI exposure, when considering the presence or absence of XAI exposure, we do not include a Probability node in Supplementary Figure 6C. However, this Probability implicitly interacts with not only the Subsequent Confidence but also the XAI components (for example, Supplementary Figures 6A and 6B). Hence, between the AI-only and XAI cohorts, one possible interpretation for the differences is as follows. First, for AI-correct subset, when images were subjectively easier, viewing the model Probability may have reinforced participants’ initial intuitions. This leads to a significant positive effect on the path Prior Confidence → Subsequent Confidence → Follow AI. Second, for AI-incorrect subset, when images were subjectively harder, the participants may be unable to fully judge the model Probability and/or how this Probability can interact with the XAI components. Therefore, statistically they were not more or less confident after seeing the model Probability and the XAI outputs. Thus, in this case, Subsequent Confidence is not a mediator between their Prior Confidence and their decision to Follow AI.

*Mediation analysis of XAI Presence on user-hard subset*

Following the rationale in section “*Mediation analysis on AI-correct user-hard image subset”*, we now estimate the XAI Presence effect on Follow AI in the AI-correct user-hard subset based on the diagram in Supplementary Figure 6C.

There were no significant direct or indirect effects (through Subsequent Confidence) from XAI Presence on Follow AI, supporting previous findings where XAI components were not yet convincing to the participants (Supplementary Table 9B). However, the direct effect Prior Confidence → Follow AI was negatively significant (Supplementary Table 10B Row 1), indicating that the low confidence participants are more likely to Follow AI, and vice versa. However, the indirect effect Prior Confidence → Subsequent Confidence → Follow AI was positively significant (Supplementary Table 10B Row 2). As discussed in section “*Mediation analysis on XAI Presence*“, although not included in this analysis, the model Probability was likely implicitly interacting with the path Prior Confidence → Subsequent Confidence → Follow AI. Hence, participants feel more confident, likely due to how the model Probability was interacting with their initial intuitions.
