## Supplemental Figures for "Application of deep learning and explainable AI-supported medical decision-making for facial phenotyping in genetic syndromes"

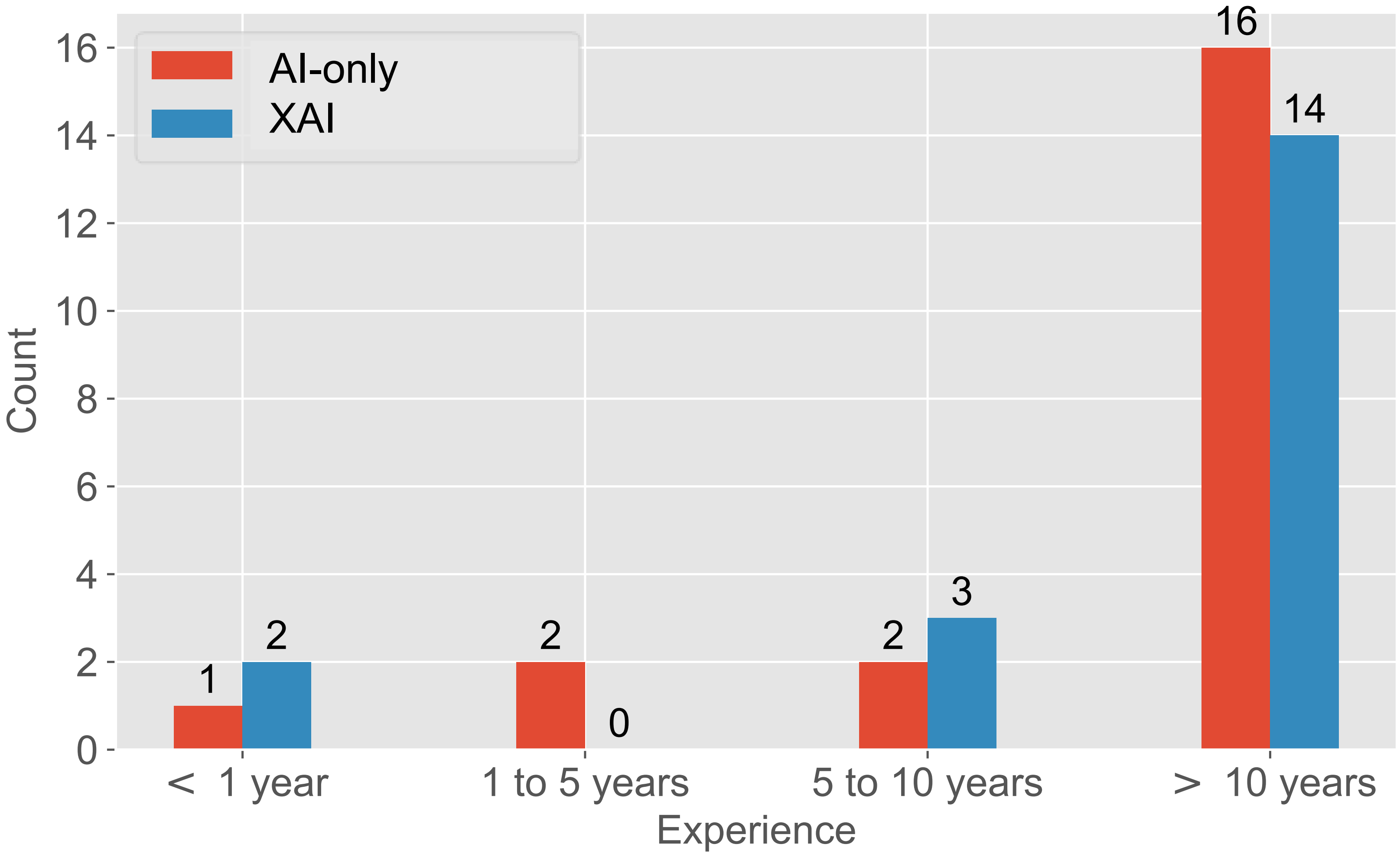


**Supplementary Figure 1**: Participants' years of experience in rare genetic diseases.

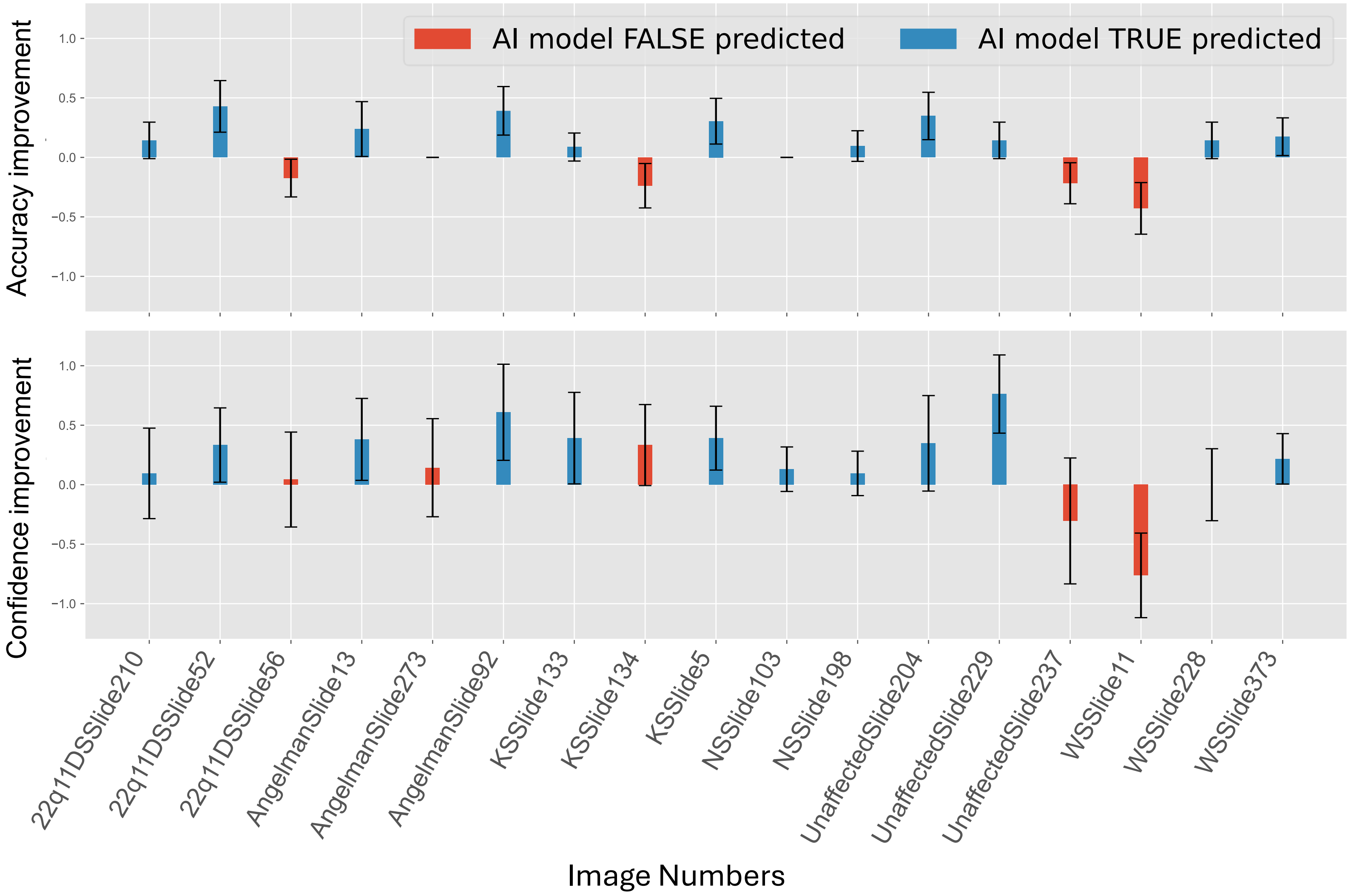


**Supplementary Figure 2**: Accuracy and confidence improvement in the AI-only group, viewed separately for each image (17 images in total). Cases where the AI model's classification was incorrect (false) are marked in red. The only intervention provided was the AI model's prediction probability.

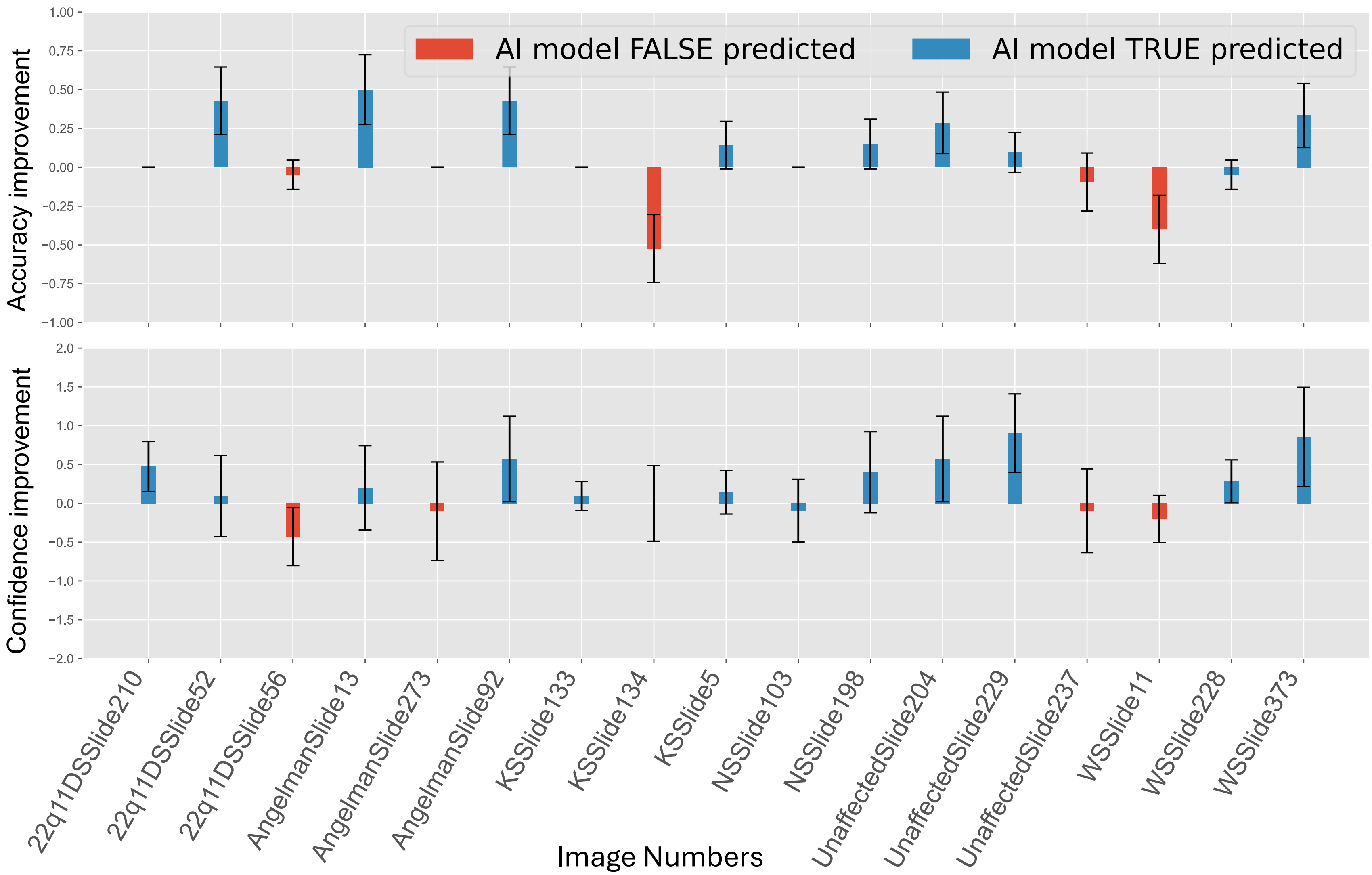


**Supplementary Figure 3**: Accuracy and confidence improvement in the XAI group, viewed separately for each image (17 images in total). Cases where the AI model's classification was incorrect (false) are marked in red. The intervention provided was the AI model's prediction probability, saliency maps, and region relevance scores.

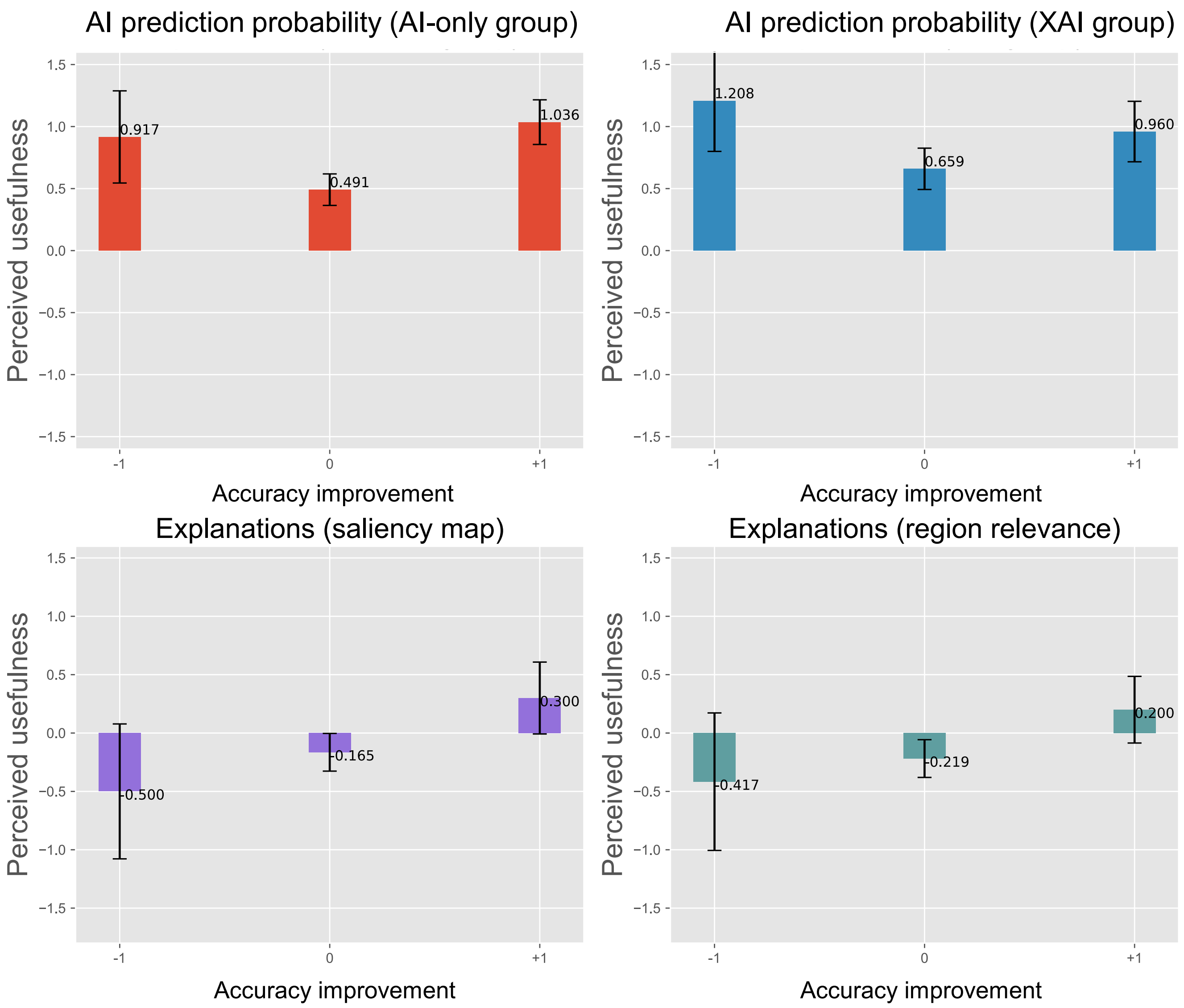


**Supplementary Figure 4**: The relationship between accuracy improvement and participants' perceived usefulness ratings of each AI/XAI component. AI prediction probability in the AI-only and XAI groups (top), and explanations: saliency map (bottom left) and region relevance scores (bottom right).

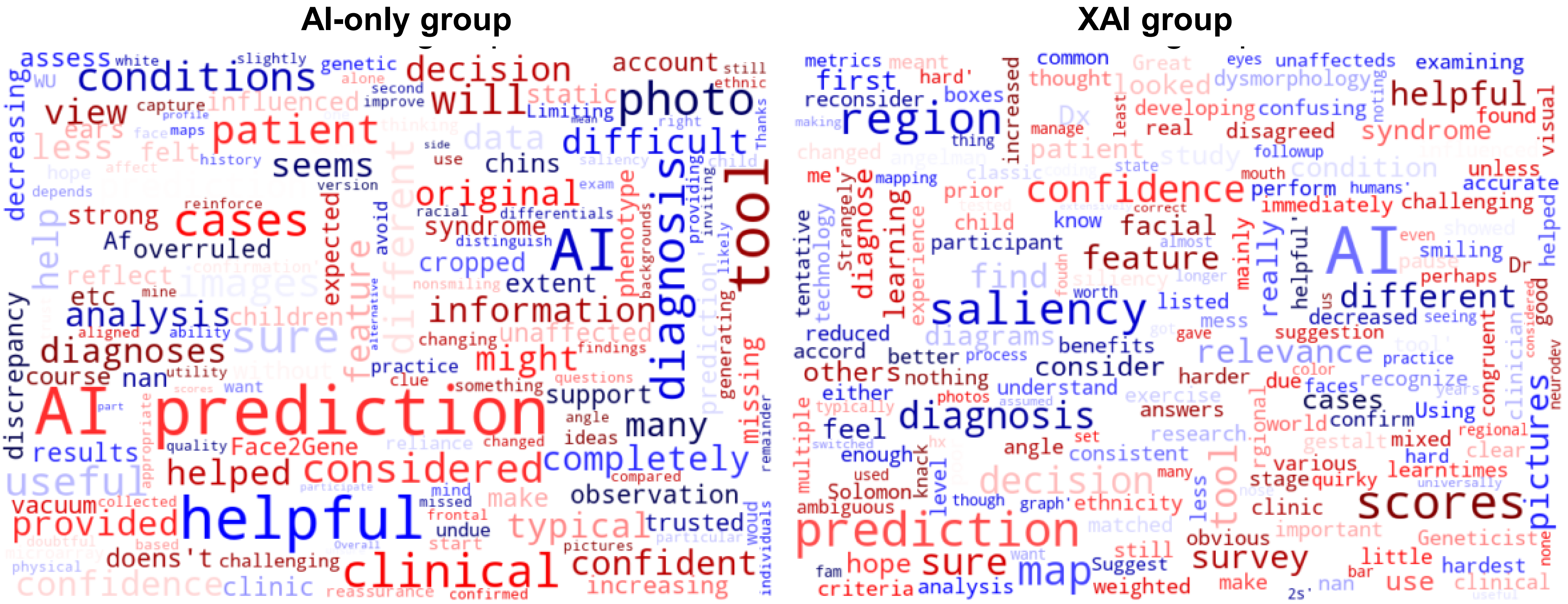


**Supplementary Figure 5**: Word cloud representations of free-text feedback from participants in the AI-only and XAI groups, illustrating the most frequently used terms.

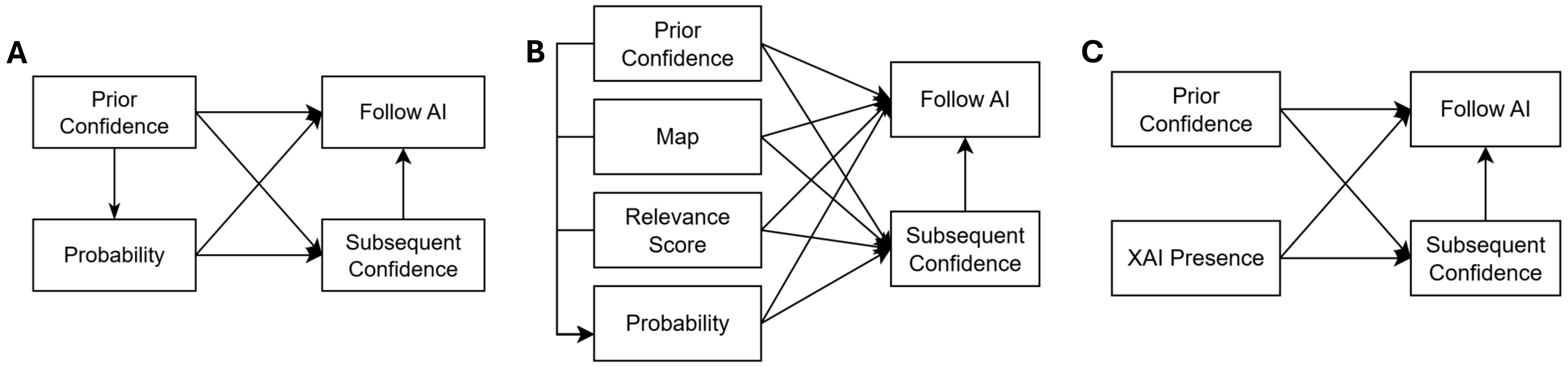


**Supplementary Figure 6. Mediation model diagrams.** Left: Diagram for AI-only cohort AI-correct and –incorrect subsets. Center: Diagram for XAI cohort AI-correct and –incorrect subsets. Right: Diagram modeling XAI exposure as an input; here, the entire undivided dataset is used for the mediation analysis. Prior Confidence is the participant initial confidence (e.g., Likert scale from -2 to 2). Subsequent Confidence is how users feel after being exposed to the model. Follow AI denotes whether participants follow the model prediction. Other abbreviated terms are as follows: Probability refers to the model prediction; Map refers to the saliency map; Relevance Score refers to the region relevance score histogram.
