## Supplemental Tables for "Application of deep learning and explainable AI-supported medical decision-making for facial phenotyping in genetic syndromes"

**Supplementary Table 1**: Participants' affiliated institutions in Baseline and XAI groups.

| Institution type | Baseline group | XAI group |
| --- | --- | --- |
| Academic medical or research center | 17 | 19 |
| Community based hospital | 4 | 1 |
| Molecular diagnostic company | 1 | 1 |
| Other (Government Research Institute) | 1 | 0 |

**Supplementary Table 2A**: Users’ Qualitative Feedback on AI and XAI Support

**AI-only group**

AI overruled my clinical when there was a discrepancy. This doesn’t reflect how we make diagnoses in clinic where we have much more information to take into account rather than just an image.

Main observation was that it was difficult to assess some cases with static and/or cropped images (missing ears, chins, etc.).

AI prediction was a strong support to my original prediction

Considered the AI prediction to some extent when I felt sure it was a syndrome rather than unaffected.

The AI results mostly influenced me by decreasing or increasing my confidence in the a diagnosis.

I am not sure how much the AI predictions helped. Just like Face2Gene I wasn’t sure I trusted AI that much. Of course this is in vacuum without any clinical information. In AfAm children AI might be more helpful as I think the typical phenotype can be different than expected and challenging. AI can be helpful in generating ideas but I woud hope we avoid an undue reliance in clinical practice This tool is not helpful. Limiting to 5 conditions when there are many more genetic conditions. Typical WU will start with exome or microarray which will capture most of these and other conditions.

I mostly use it if I have no clue what the child could have, or if I am thinking something but want confirmation

AI prediction was more useful in providing more confidence (reassurance) than completely changing mind (help to distinguish between differentials). I completely missed the saliency maps on my view. for the original questions - the utility of this depends on the particular patient (remainder of clinical history and other physical exam findings).

Useful tool. Some cases I was still doubtful

When AI prediction aligned with my diagnosis, it was helpful. However, when it was not, I did not trust the AI prediction over mine. 25In the second version of the photo (when the AI prediction answer was provided), I changed my answer to the one that the prediction provided. However, there were a few I am less confident on the prediction’s ability. I also think a nonsmiling face with the right angle is most appropriate for analysis, and many of these photos did not have that feature which could affect the analysis of the tool.

When I wasn’t sure based on the photo alone, the AI prediction was helpful.

Some pictures were difficult because of the quality of the image or having only the frontal view. In some cases, I was more confident in my decision and less so in the AI prediction tool. In some cases, the AI tool helped to reinforce my decision. It seems like having more images and more data on patients with confirmed diagnoses would help improve the tool. It's also helpful to have images of patients with different racial and ethnic backgrounds because they might have slightly different features of a diagnosis compared to white individuals where likely most of the data has been collected. Thanks for inviting me to participate. Overall, it seems like the tool will be useful!

I considered AI when I had alternative diagnosis or am otherwise unsure. For the most part, the scores did not mean much to me. For the photos, having a side profile would have been very helpful.

**XAI group**

I did not find the saliency maps or rgional scores helpful. The gestalt is what is important.

I don’t know how the AI does with ethnicity

When my decision was congruent with the AI tool it increased my confidence but when not

it decreased my confidence on a diagnosis. The feature of the siliency and prediction were

good but not the tool that weighted the facial regions. I found that angelman syndrome was

the hardest for me to diagnose and I was not very sure of the AI tools confidence. I hope I

can get the answers and learn from this.

Did not consider AI prediction. Some cases harder than others due to smiling child or at different angle. I had little use of the saliency/region relevance scores in decision, unless they region scores are consistent enough to be listed at common criteria.

Using pictures is more challenging that examining a patient in the clinic. I feel some pictures mixed different features of the conditions which was confusing. I feel that AI is still developing. A prior study by Dr. Solomon showed that Geneticist perform better than AI for dysmorphology.

I did not find the map and region scores at all helpful.

not sure how to use this tool

My first thought is I hope I don’t mess up your survey by being a poor research participant I really did not understand the AI diagrams and pictures either I recognize the condition immediately stage with that diagnosis or if the AI tool disagreed I reduced my confidence level and if this was the real world it would make me pause and reconsider am I really sure this is the diagnosis which as for the various boxes and diagrams they meant nothing to me

The AI prediction matched the clinical diagnosis in most of the cases.

saliency maps has no clear benefits to me

Great exercise on how much more accurate facial analysis technology is than an experience clinician (me)

I did not find the saliency or region relevance scores helpful

I did consider it multiple times and changed my answer. The scores, mainly the predictions, influenced my decision.

Some diagnoses obvious and my visual Dx and AI were in accord. For those where I was less sure the AI Dx helped confirm my tentative suggestion. Strangely the unaffecteds were hard

This is a quirky survey. Some faces were classic, others ambiguous, but perhaps that was your study. It was hard to get the knack of the AI metrics. Suggest you do a followup survey since there is a learning process for us humans

As I got more used to the set up the first thing I looked at was the prediction then the mapping, and least helpful was the regional bar graph

After seeing the color coding of eyes / nose / mouth, I foudn that I no longer looked at the saliency maps. Also, it's worth noting that even though I've been in practice for years, I typically manage patients with a different syndrome (none tested on).

would want state of neurodev features and fam hx/photos before I gave myself many 2s

I almost universally switched to the AI prediction. The saliency maps and region relevance scores were more useful for learning (I assumed the AI was correct) than for making decision.

I considered the AI prediction extensively.

**Supplementary Table 2B:** Expert commentary

Expert commentary 1

“*The saliency map, particularly in the eye region, helps emphasize the downslanting palpebral fissures (the angle of the eyes), which is a very characteristic/classic sign of Noonan syndrome. When I look at the saliency image, the first thing I notice is the saliency map showing the ways the eyes are positioned.”*

Expert commentary 2

*“I noticed that the eyebrows are noticeably highlighted. Although the eyebrows of people with Kabuki syndrome can have a characteristic appearance, this fact may not be very well known to clinicians (including the survey respondents), and this may have been distracting or led respondents to doubt the diagnosis of Kabuki syndrome. [...] The image is rather difficult, I feel. It does have some features that I feel are indeed reminiscent of 22q11.2 deletion syndrome, such as the shape of the nose, including the nasal bridge. More broadly, 22q11.2 is often quite difficult to identify based on facial findings unless clinicians are very experienced with this condition.''*

**Supplementary Table 3.** How participant responses are considered to Follow AI
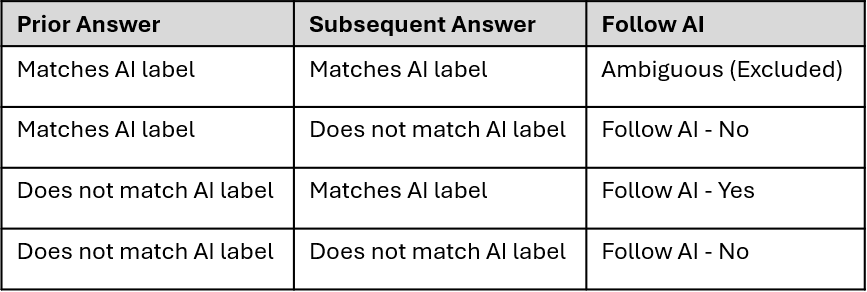

For example, in Figure 5A, the AI classification label is "Noonan syndrome". If the participant's Initial Answer is "unaffected" and the participant's Subsequent Answer is "Noonan syndrome", the participant is considered to Follow AI. If the participant's Prior Answer and the Subsequent Answer are both "Noonan syndrome", they are considered to be Ambiguous. These Ambiguous responses are removed from the mediation analysis.

**Supplementary Table 4A-D.** Mediation analysis in AI-only AI-correct and –incorrect subset

| **Supp. Table 4A. AI-only AI-correct subset** | | | |
| --- | --- | --- | --- |
| **Regression coefficients from mediation equations** | | | |
|  | **Estimate** | **Std.Err** | **P-value** |
| **Probability ~** |  |  |  |
| Prior Confidence | 0.383 | 0.100 | 0 |
| **Subsequent Confidence ~** |  |  |  |
| Prior Confidence | 0.178 | 0.076 | 0.019 |
| Probability | 0.66 | 0.080 | 0 |
| **Follow AI~** |  |  |  |
| Prior Confidence | -1.242 | 0.508 | 0.015 |
| Subsequent Confidence | 0.2812 | 0.617 | 0.649 |
| Probability | 1.5234 | 0.582 | 0.009 |

| **Supp. Table 4B. AI-only AI-correct subset** | | |  | |
| --- | --- | --- | --- | --- |
| **Mediation path** | **Direct Effect** | | **Indirect Effect** | |
|  | **Estimate** | **95% Conf.** | **Estimate** | **95% Conf.** |
| PC🡪FAI | -1.242 | [-2.238, -0.246] * |  |  |
| PC🡪SC🡪FAI |  |  | 0.050 | [-0.155, 0.255] |
| PC🡪Prob🡪FAI |  |  | 0.583 | [0.010, 1.155] * |
| PC🡪Prob🡪SC🡪FAI |  |  | 0.071 | [-0.234, 0.376] |
| **Total** |  |  | -0.539 | [-1.477, 0.400] |

| **Supp. Table 4C. AI-only AI-incorrect subset** | | | |
| --- | --- | --- | --- |
| **Regression coefficients from mediation equations** | | | |
|  | **Estimate** | **Std.Err** | **P-value** |
| **Probability ~** |  |  |  |
| Prior Confidence | 0.187 | 0.115 | 0.104 |
| **Subsequent Confidence ~** |  |  |  |
| Prior Confidence | 0.397 | 0.079 | 0 |
| Probability | 0.248 | 0.072 | 0.001 |
| **Follow AI~** |  |  |  |
| Prior Confidence | 0.032 | 0.303 | 0.917 |
| Subsequent Confidence | -0.914 | 0.477 | 0.055 |
| Probability | 1.786 | 0.439 | 4.71E-05 |

| **Supp. Table 4D. AI-only AI-incorrect subset** | | |  | |
| --- | --- | --- | --- | --- |
| **Mediation path** | **Direct Effect** | | **Indirect Effect** | |
|  | **Estimate** | **95% Conf.** | **Estimate** | **95% Conf.** |
| PC🡪FAI | 0.032 | [-0.562, 0.626] |  |  |
| PC🡪SC🡪FAI |  |  | -0.363 | [-0.801, 0.075] |
| PC🡪Prob🡪FAI |  |  | 0.333 | [-0.167, 0.833] |
| PC🡪Prob🡪SC🡪FAI |  |  | -0.042 | [-0.122, 0.038] |
| **Total** |  |  | -0.04 | [-0.8, 0.72] |

Equations below are used for mediation analysis based on Supplementary Figure 6A. Each epsilon is random noise. Intercept terms are included in each equation but not shown here for brevity. The last equation is fitted with logistic regression, instead of linear regression, because of the binary nature of Follow AI. Regression coefficients are shown on the top tables. Abbreviations used are PC for Prior Confidence, SC for Subsequent Confidence, PR is Probability of model prediction, FA is Follow AI.

$$Pr=\beta_{PC,PR}PC+ \epsilon_{PR}$$

$$SC={\beta_{PC,SC}PC+ \beta}_{Pr,SC}PR+\epsilon_{SC}$$

$$\log\frac{FA}{1-FA}= \beta_{PC,FA}PC+\beta_{PR,FA}PR+ \beta_{SC,FA}SC+\epsilon_{FA}$$

Direct and indirect mediation effects are on the bottom table. The 95% confidence intervals for the indirect effects are computed via bootstrap (* denotes statistical significance). For example, the direct effect PC 🡪 FA is simply $\beta_{PC,FA}.$The indirect effect PC 🡪 PR 🡪 FA is computed as the product $\beta_{PC,PR}\beta_{PR,FA}$. Similarly indirect effect PC 🡪PR 🡪 SC 🡪 FA is $\beta_{PC,PR}\beta_{PR,SC}\beta_{SC,FA}$. The total effect of PC on FA is computed by adding the effects of all the paths.

**Supplementary Table 5A-D.** Mediation analysis in XAI AI-correct and –incorrect subset.

| **Supp. Table 5A. XAI AI-correct** | | | |
| --- | --- | --- | --- |
| **Regression coefficients from mediation equations** | | | |
|  | **Estimate** | **Std.Err** | **P-value** |
| **Map ~** |  |  |  |
| Prior Confidence | -0.160 | 0.145 | 0.269 |
| **Relevance Score ~** |  |  |  |
| Prior Confidence | -0.222 | 0.130 | 0.087 |
| **Probability ~** |  |  |  |
| Prior Confidence | -0.210 | 0.131 | 0.110 |
| **Subsequent Confidence ~** |  |  |  |
| Prior Confidence | 0.130 | 0.110 | 0.237 |
| Map | 0.361 | 0.087 | 0.000 |
| Relevance Score | -0.276 | 0.097 | 0.004 |
| Probability | 0.323 | 0.096 | 0.001 |
| **Follow AI~** |  |  |  |
| Prior Confidence | 0.045 | 0.335 | 0.894 |
| Subsequent Confidence | -0.121 | 0.407 | 0.767 |
| Map | 0.500 | 0.701 | 0.476 |
| Relevance Score | -0.532 | 0.819 | 0.516 |
| Probability | 1.629 | 0.542 | 0.003 |

| **Supp. Table 5B. XAI AI-correct** | | |  | |
| --- | --- | --- | --- | --- |
| **Mediation path** | **Direct Effect** | | **Indirect Effect** | |
|  | **Estimate** | **95% Conf.** | **Estimate** | **95% Conf.** |
| PC 🡪 FAI | 0.045 | [-0.612, 0.702] |  |  |
| PC 🡪 SC🡪 FAI |  |  | -0.016 | [-0.107, 0.076] |
| PC 🡪 Map 🡪 FAI |  |  | -0.080 | [-0.3, 0.139] |
| PC 🡪 RS 🡪 FAI |  |  | 0.118 | [-0.213, 0.45] |
| PC 🡪 Prob 🡪 FAI |  |  | -0.342 | [-0.742, 0.058] |
| PC 🡪 Map 🡪 SC 🡪 FAI |  |  | 0.007 | [-0.024, 0.038] |
| PC 🡪 RS 🡪SC 🡪 FAI |  |  | -0.007 | [-0.039, 0.024] |
| PC 🡪 Prob 🡪 SC 🡪 FAI |  |  | 0.008 | [-0.028, 0.045] |
| **Total** |  |  | -0.267 | [-1.372, 0.838] |

| **Supp. Table 5C. XAI AI-incorrect** | | | |
| --- | --- | --- | --- |
| **Regression coefficients from mediation equations** | | | |
|  | **Estimate** | **Std.Err** | **P-value** |
| **Map ~** |  |  |  |
| Prior Confidence | -0.298 | 0.121 | 0.014 |
| **Relevance Score ~** |  |  |  |
| Prior Confidence | -0.231 | 0.127 | 0.070 |
| **Probability ~** |  |  |  |
| Prior Confidence | -0.126 | 0.149 | 0.399 |
| **Subsequent Confidence ~** |  |  |  |
| Prior Confidence | 0.500 | 0.093 | 0.000 |
| Map | -0.029 | 0.080 | 0.721 |
| Relevance Score | -0.018 | 0.076 | 0.810 |
| Probability | 0.188 | 0.065 | 0.004 |
| **Follow AI~** |  |  |  |
| Prior Confidence | -0.902 | 0.481 | 0.061 |
| Subsequent Confidence | -0.492 | 0.522 | 0.346 |
| Map | -2.755 | 3.426 | 0.421 |
| Relevance Score | 2.720 | 3.429 | 0.428 |
| Probability | 2.163 | 0.537 | 0.000 |

| **Supp. Table 5D. XAI AI-incorrect** | | |  | |
| --- | --- | --- | --- | --- |
| **Mediation path** | **Direct Effect** | | **Indirect Effect** | |
|  | **Estimate** | **95% Conf.** | **Estimate** | **95% Conf.** |
| PC 🡪 FAI | -0.902 | [-1.845, 0.041] |  |  |
| PC 🡪 SC🡪 FAI |  |  | -0.246 | [-0.774, 0.282] |
| PC 🡪 Map 🡪 FAI |  |  | 0.822 | [-0.035, 1.679] |
| PC 🡪 RS 🡪 FAI |  |  | -0.627 | [-1.433, 0.178] |
| PC 🡪 Prob 🡪 FAI |  |  | -0.272 | [-1.005, 0.46] |
| PC 🡪 Map 🡪 SC 🡪 FAI |  |  | -0.004 | [-0.048, 0.039] |
| PC 🡪 RS 🡪SC 🡪 FAI |  |  | -0.002 | [-0.031, 0.027] |
| PC 🡪 Prob 🡪 SC 🡪 FAI |  |  | 0.012 | [-0.02, 0.044] |
| **Total** |  |  | -1.220 | [-2.915, 0.476] |

Equations below are used for mediation analysis based on Supplementary Figure 6B. Each epsilon is random noise. Intercept terms are included in each equation but not shown here for brevity. The last equation is fitted with logistic regression, instead of linear regression, because of the binary nature of Follow AI. Regression coefficients are shown on the top tables. Abbreviations for PC, SC, PR, and FA are as defined in Supplementary Table 4; here, Map is Saliency Map, RS is region relevance score histogram.

$$Map=\beta_{PC,Map}PC+ \epsilon_{Map}$$

$$RS=\beta_{PC,RS}PC+ \epsilon_{RS}$$

$$PR=\beta_{PC,PR}PC+ \epsilon_{PR}$$

$$SC={\beta_{PC,SC}PC+\beta}_{Map,SC}Map+{\beta_{RS,SC}RS+\beta}_{PR,SC}PR+ \epsilon_{SC}$$

$$\log\frac{FA}{1-FA}= \beta_{PC,FA}PC+{\beta_{Map,FA}Map+\beta_{RS,FA}RS+\beta}_{Prob,FA}PR+ \beta_{SC,FA}SC+\epsilon_{FA}$$

Direct and indirect mediation effects are on the bottom table. The 95% confidence intervals for the indirect effects are computed via bootstrap (* denotes statistical significance).

**Supplementary Table 6A-D.** Mediation analysis of XAI Presence AI-correct and –incorrect subset.

| **Supp. Table 6A. XAI Presence AI-correct subset** | | | |
| --- | --- | --- | --- |
| **Regression coefficients from mediation equations** | | | |
|  | **Estimate** | **Std.Err** | **P-value** |
| Subsequent Confidence ~ |  |  |  |
| Prior Confidence | 0.247 | 0.077 | 0.001 |
| XAI Presence | -0.012 | 0.147 | 0.933 |
| Follow AI~ |  |  |  |
| Prior Confidence | -0.526 | 0.245 | 0.032 |
| XAI Presence | -0.119 | 0.394 | 0.763 |
| Subsequent Confidence | 0.715 | 0.236 | 0.002 |

| **Supp. Table 6B. XAI Presence AI-correct subset** | | |  | |
| --- | --- | --- | --- | --- |
| **Mediation path** | **Direct Effect** | | **Indirect Effect** | |
|  | **Estimate** | **95% Conf.** | **Estimate** | **95% Conf.** |
| PC -> FAI | -0.526 | [-1.006, -0.046] * |  |  |
| PC -> SC -> FAI |  |  | 0.177 | [0.026, 0.328]* |
| **Total** |  |  | -0.349 | [-0.916, 0.218] |
| XAI Presence -> FAI | -0.119 | [-0.891, 0.663] |  |  |
| XAI Presence -> SC -> FAI |  |  | -0.009 | [-0.205, 0.188] |
| **Total** |  |  | -0.128 | [-0.967, 0.712] |

| **Supp. Table 6C. XAI Presence AI-Incorrect subset** | | | |
| --- | --- | --- | --- |
| **Regression coefficients from mediation equations** | | | |
|  | **Estimate** | **Std.Err** | **P-value** |
| Subsequent Confidence ~ |  |  |  |
| Prior Confidence | 0.466 | 0.062 | 0.000 |
| XAI Presence | 0.098 | 0.128 | 0.441 |
| Follow AI~ |  |  |  |
| Prior Confidence | -0.365 | 0.181 | 0.044 |
| XAI Presence | 0.360 | 0.315 | 0.252 |
| Subsequent Confidence | 0.314 | 0.193 | 0.104 |

| **Supp. Table 6D. XAI Presence AI-incorrect subset** | | |  | |
| --- | --- | --- | --- | --- |
| **Mediation path** | **Direct Effect** | | **Indirect Effect** | |
|  | **Estimate** | **95% Conf.** | **Estimate** | **95% Conf.** |
| PC -> FAI | -0.365 | [-0.72, -0.01] * |  |  |
| PC -> SC -> FAI |  |  | 0.146 | [-0.041, 0.333] |
| **Total** |  |  | -0.219 | [-0.538, 0.101] |
| XAI Presence -> FAI | 0.360 | [-0.977, 0.257] |  |  |
| XAI Presence -> SC -> FAI |  |  | 0.031 | [-0.05, 0.112] |
| **Total** |  |  | 0.391 | [-0.235, 1.017] |

Equations below are used for mediation analysis based on Supplementary Figure 6C. Each epsilon is random noise. Intercept terms are included in each equation but not shown here for brevity. The last equation is fitted with logistic regression, instead of linear regression, because of the binary nature of Follow AI. Regression coefficients are shown on the top tables. Abbreviations for PC, SC, and FA are defined in Supplementary Table 4. Abbreviations for Map and RS are defined in Supplementary Table 5. XAI Presence is abbreviated to XAI.

$$SC=\beta_{PC, SC}PC+\beta_{XAI, SC}XAI+\epsilon_{SC}$$

$$\log\frac{FA}{1-FA}= \beta_{PC,FA}PC+\beta_{XAI,FA}XAI+ \beta_{SC,FA}SC+\epsilon_{FA}$$

Direct and indirect mediation effects are on the bottom table. The 95% confidence intervals for the indirect effects are computed via bootstrap (* denotes statistical significance).

**Supplementary Table 7.** Prior accuracy percentage of each image averaging over each cohort and on the whole dataset. Images with an average prior accuracy over 75% are considered user-easy. Likewise, images with a prior accuracy under 75% are considered user-hard. Image number 9 was removed due to recording error.

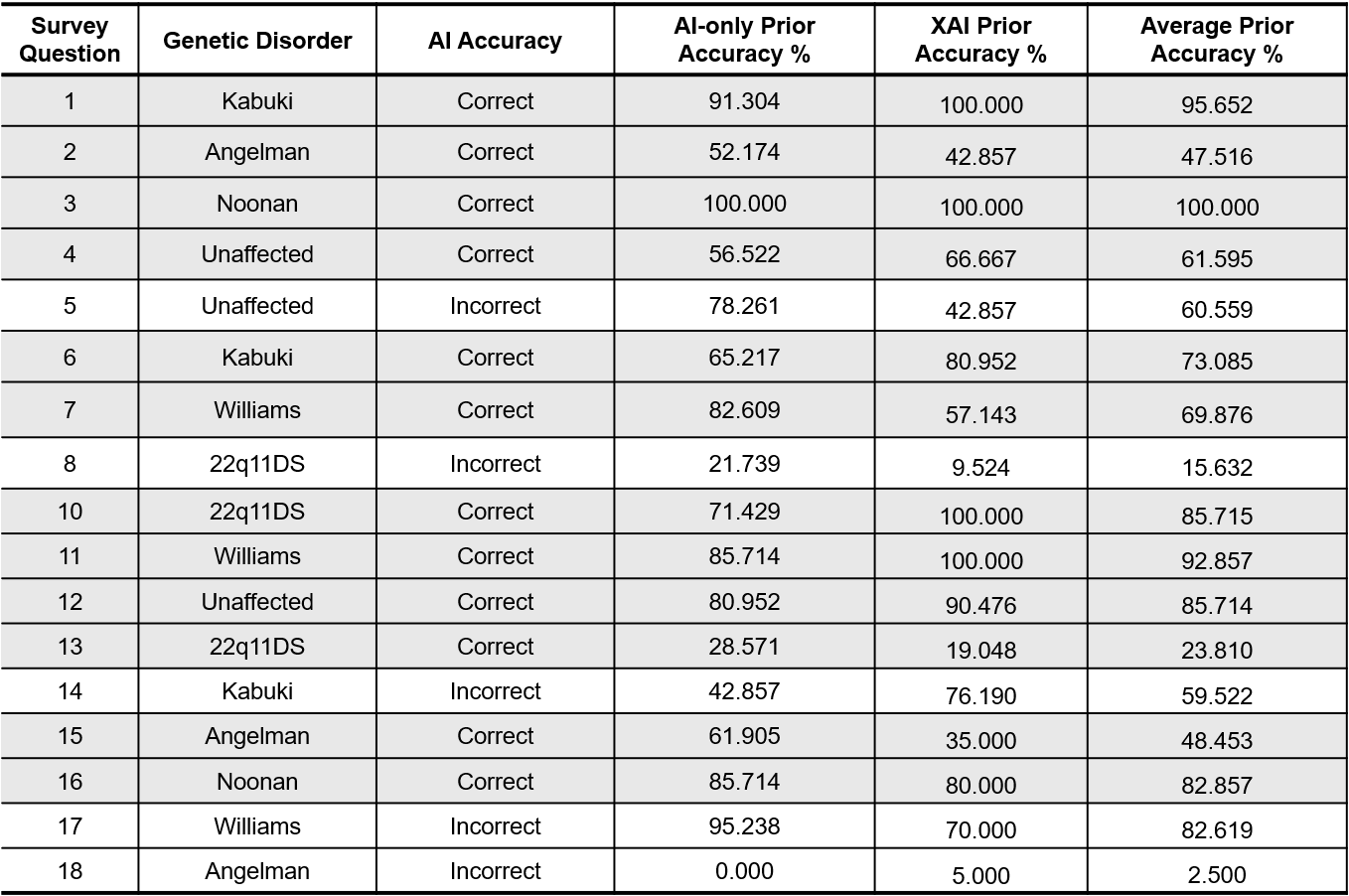

**Supplementary Table 8A-B.** Mediation analysis of AI-correct user-hard subset on AI-only cohort. Equations used for mediation analysis and abbreviations for PC, SC, PR, and FA are shown in Supplementary Table 4. Equations are based on Supplementary Figure 6A.

| **Supp. Table 8A. AI-only AI-correct User-hard subset** | | | |
| --- | --- | --- | --- |
| **Regression coefficients from mediation equations** | | | |
|  | **Estimate** | **Std.Err** | **P-value** |
| **Probability ~** |  |  |  |
| Prior Confidence | 0.448 | 0.120 | 0.000 |
| **Subsequent Confidence ~** |  |  |  |
| Prior Confidence | 0.222 | 0.093 | 0.017 |
| Probability | 0.610 | 0.092 | 0.000 |
| **Follow AI~** |  |  |  |
| Prior Confidence | -2.430 | 1.078 | 0.024 |
| Subsequent Confidence | -0.670 | 1.068 | 0.530 |
| Probability | 3.158 | 1.253 | 0.012 |

| **Supp. Table 8B. AI-only AI-correct User-hard subset** | | | | |
| --- | --- | --- | --- | --- |
| **Mediation path** | **Direct Effect** | | **Indirect Effect** | |
|  | **Estimate** | **95% Conf.** | **Estimate** | **95% Conf.** |
| PC🡪FAI | -2.430 | [-3.429, -1.431] * |  |  |
| PC🡪SC🡪FAI |  |  | -0.149 | [-0.396, 0.098] |
| PC🡪Prob🡪FAI |  |  | 1.413 | [0.668, 2.159] * |
| PC🡪Prob🡪SC🡪FAI |  |  | -0.183 | [-0.471, 0.105] |
| **Total** |  |  | -1.348 | [-2.424, -0.273] |

**Supplementary Table 9A-B. Mediation analysis of AI-correct user-hard subset on XAI cohort.** Equations used for mediation analysis and abbreviations for PC, SC, Map, PR, RS, and FA are shown in Supplementary Table 5. Equations are based on Supplementary Figure 6B.

| **Supp. Table 9A. XAI-only AI-correct User-hard subset** | | | |
| --- | --- | --- | --- |
| **Regression coefficients from mediation equations** | | | |
|  | **Estimate** | **Std.Err** | **P-value** |
| **Map ~** |  |  |  |
| Prior Confidence | -0.124 | 0.149 | 0.404 |
| **Relevance Score ~** |  |  |  |
| Prior Confidence | -0.192 | 0.131 | 0.142 |
| **Probability ~** |  |  |  |
| Prior Confidence | -0.139 | 0.134 | 0.302 |
| **Subsequent Confidence ~** |  |  |  |
| Prior Confidence | 0.117 | 0.106 | 0.270 |
| Map | 0.367 | 0.088 | 0.000 |
| Relevance Score | -0.233 | 0.099 | 0.019 |
| Probability | 0.343 | 0.097 | 0.000 |
| **Follow AI~** |  |  |  |
| Prior Confidence | 0.025 | 0.441 | 0.945 |
| Subsequent Confidence | 0.115 | 0.358 | 0.797 |
| Map | 0.411 | 0.536 | 0.592 |
| Relevance Score | -0.279 | -0.314 | 0.754 |
| Probability | 1.727 | 2.844 | 0.004 |

| **Supp. Table 9B. XAI-only AI-correct User-hard subset** | | | | |
| --- | --- | --- | --- | --- |
| **Mediation path** | **Direct Effect** | | **Indirect Effect** | |
|  | **Estimate** | **95% Conf.** | **Estimate** | **95% Conf.** |
| PC 🡪 FAI | 0.025 | [-0.839, 0.889] |  |  |
| PC 🡪 SC🡪 FAI |  |  | 0.013 | [-0.072, 0.099] |
| PC 🡪 Map 🡪 FAI |  |  | -0.051 | [-0.259, 0.157] |
| PC 🡪 RS 🡪 FAI |  |  | 0.054 | [-0.263, 0.371] |
| PC 🡪 Prob 🡪 FAI |  |  | -0.24 | [-0.593, 0.114] |
| PC 🡪 Map 🡪 SC 🡪 FAI |  |  | -0.005 | [-0.031, 0.021] |
| PC 🡪 RS 🡪SC 🡪 FAI |  |  | 0.005 | [-0.02, 0.03] |
| PC 🡪 Prob 🡪 SC 🡪 FAI |  |  | -0.005 | [-0.029, 0.018] |
| **Total** |  |  | -0.204 | [-1.391, 0.982] |

**Supplementary Table 10.** Mediation analysis of XAI Presence in AI-correct user-hard subset. Equations used for mediation analysis and abbreviations for PC, SC, FA, and XAI are shown in Supplementary Table 6. Equations are based on Supplementary Figure 6C.

| **Supp. Table 10A. XAI Presence AI-correct User-hard** | | | |
| --- | --- | --- | --- |
| **Regression coefficients from mediation equations** | | | |
|  | **Estimate** | **Std.Err** | **P-value** |
| Subsequent Confidence ~ |  |  |  |
| Prior Confidence | 0.247 | 0.085 | 0.004 |
| XAI Presence | -0.027 | 0.160 | 0.868 |
| Follow AI~ |  |  |  |
| Prior Confidence | -0.593 | 0.282 | 0.036 |
| XAI Presence | -0.145 | 0.438 | 0.74 |
| Subsequent Confidence | 0.766 | 0.270 | 0.005 |

| **Supp. Table 10B. XAI Presence AI-correct User-hard** | | |  | |
| --- | --- | --- | --- | --- |
| **Mediation path** | **Direct Effect** | | **Indirect Effect** | |
|  | **Estimate** | **95% Conf.** | **Estimate** | **95% Conf.** |
| PC -> FAI | -0.593 | [-0.76, -0.426] * |  |  |
| PC -> SC -> FAI |  |  | 0.189 | [1.687E-4, 0.378] * |
| **Total** |  |  | -0.404 | [-1.079, 0.272] |
| XAI Presence -> FAI | -0.145 | [-1.003, 0.713] |  |  |
| XAI Presence -> SC -> FAI |  |  | -0.02 | [0.253, 0.213] |
| **Total** |  |  | -0.166 | [-1.204, 0.873] |
